# An Analysis of PCR Ct Scores and Distributions from the ONS Community Infection Survey during the COVID Second Wave in the UK

**DOI:** 10.1101/2022.06.03.22275903

**Authors:** Keith Johnson, Steven J Hammer, Tanya Klymenko

**Affiliations:** IP Consultant for Diagnostic Testing, Thurnbichl 2, A-6345 Kössen, Austria; Data Analyst; Senior Lecturer in Biomedical Sciences, Sheffield Hallam University, City Campus, Howard Street, Sheffield, S1 1WB, UK

## Abstract

This work presents an analysis of PCR cycle threshold (Ct) scores and their distributions, i.e. the probabilities that a test is positive with a score Ct, P(Ct), derived from the survey during the second COVID wave in the UK. Their relation to gene target breakdown is exemplified. Thus a significant parameter for tracking the course of COVID in the second wave is the percentage of positive tests with Ct < 25, %Ct <25, which is obtained by plotting weekly percentiles from the survey against Ct to construct the ogive or cumulative frequency curve (CMF). The biological basis for studying this parameter is the strong correlation between %Ct < 25 and the percentage of positive tests comprising target genes ORF1ab+N and ORF1ab+N+S, or %Inf.

Furthermore, the probability distributions, obtained by differentiating the ogives, were found to be predominantly bimodal with a hot peak at Ct = 20.31+/- 4.65 and a cold peak with Ct = 32.95+/-1.11. These closely match the peaks found for the target genes ORF1ab+N, viz. Ct=18.54+/-2.31 and Ct=32.02+/-0.49 as well as in Walker et al [12]. Similar results were found in [13] and [14]. The cold peak seems likely to be associated with residue from a previous infection. The distributions for gene targets in cfvroc Pillar 2 [15,16] are also bimodal but the peaks occur at lower values of Ct. This suggests the results are machine/sample dependent and emphasises the need for calibration, if quality control in PCR testing is to be improved.

## I. Introduction

Since April 2020, ONS has been carrying out a survey of households in the UK, using qRT-PCR to monitor the prevalence of COVID-19 in the general population, the so-called ONS CIS survey [1]. A positive test is based on detecting the presence of the spike protein gene (S), the nucleocapsid protein gene (N) else the ORF1ab gene of SARS-CoV-2. Data collected includes positivity rates, incidence rates, Ct values for positive cases and gene target breakdown, all of which is split by nation with England further being broken down into regions. The ONS CIS data is published at least every 2 weeks.

This work presents an analysis of PCR cycle threshold (Ct) scores and their distributions, ie. the probabilities that a test is positive with a score Ct, P(Ct), derived from the survey during the second COVID wave in the UK and their relation to gene target breakdown. In this way it is hoped to shed light on what constitutes an infectious score. This was a contentious issue throughout Summer and Autumn 2020 with much discussion of hot and cold positives [2], while Heneghan et al stressed the importance of calibration [3].

Indeed, according to the minutes of SAGE 17th December [4] ‘… *Initial analysis from ONS Covid-19 Infection Survey (CIS) data up to 7 December showed that positive tests with high Ct values (>25) do not cluster with other positive tests (with either high or low Ct values). This suggests that they are not associated with transmission-type patterns meaning these people may be less infectious to others than those whose tests have a low Ct value*…’. Furthermore, Bullard et al demonstrated it was not possible to grow live cultures of SARS-COV2 virus from samples obtained after high cycles of PCR amplification [5]. Similar reports appeared in the Italian and Austrian press [6]. Hence it seems pertinent to investigate as a first parameter the percentage of PCR tests with a Ct score < 25.

The percentage of positive tests with Ct < 25 can be obtained from the ONS report for 29.01.2021 [7], in particular Table 6a, which lists the mean Ct and 10, 25, 50, 75, 90% percentiles for each week from 26.09.2020 to 18.01.2021. The percentiles can be used to construct an ogive or cumulative frequency curve [8]. This is done by plotting the percentiles on the x-axis against the corresponding percentage on the y-axis (blue line). The percentage of positive tests with Ct < 25 can then be read off from the graph.

**Figure 1:**
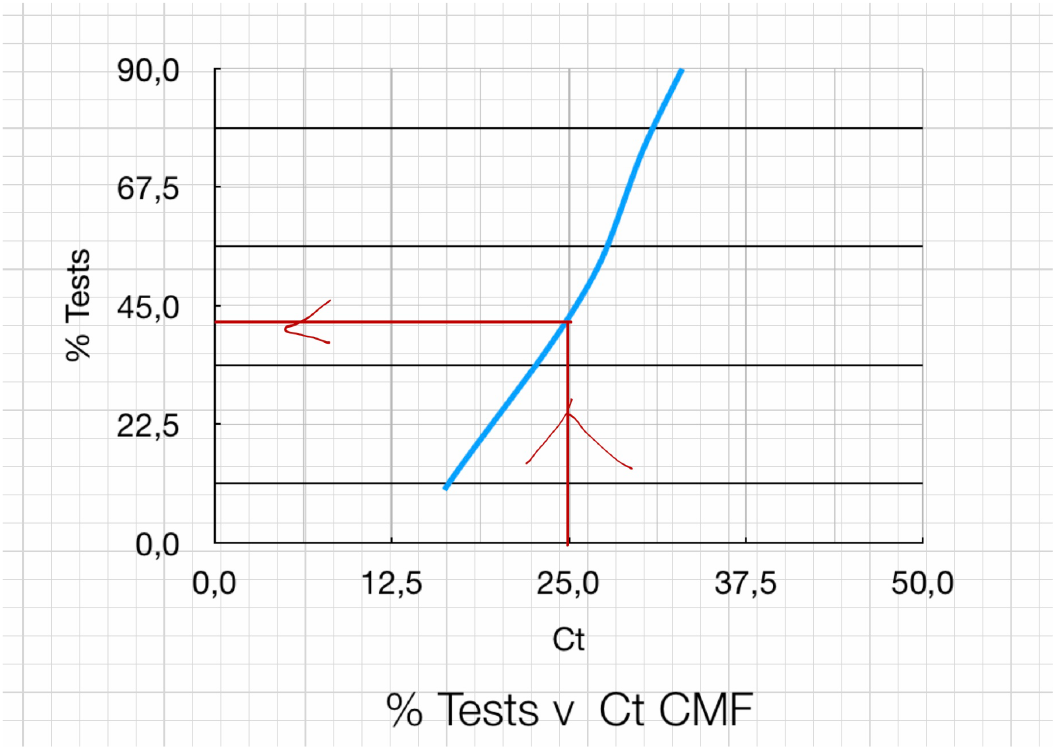
%Tests v Ct CMF

Preliminary results were obtained using Excel spreadsheets and the process was then automated using R statistical programming language [22] to provide confirmation. Full details of methods and data analysis are given in Annex I below.

## II. Infectious genes

To find a biological basis for the importance of this percentage, regional %Ct < 25 is compared with the corresponding percentage of positive tests comprising target genes ORF1ab and ORF1ab+N+S, or %Inf in what follows. Previous unpublished reports in our lab [9] brought out the different behaviours in the North (YH, NW, NE), Midlands (WM, EM, SW) and LSE (L, SE, EE). Consequently, the data were examined to see whether the same trend could be identified here too. The results are detailed in Annex I. The panel for the North is shown below:

**Figure 2:**
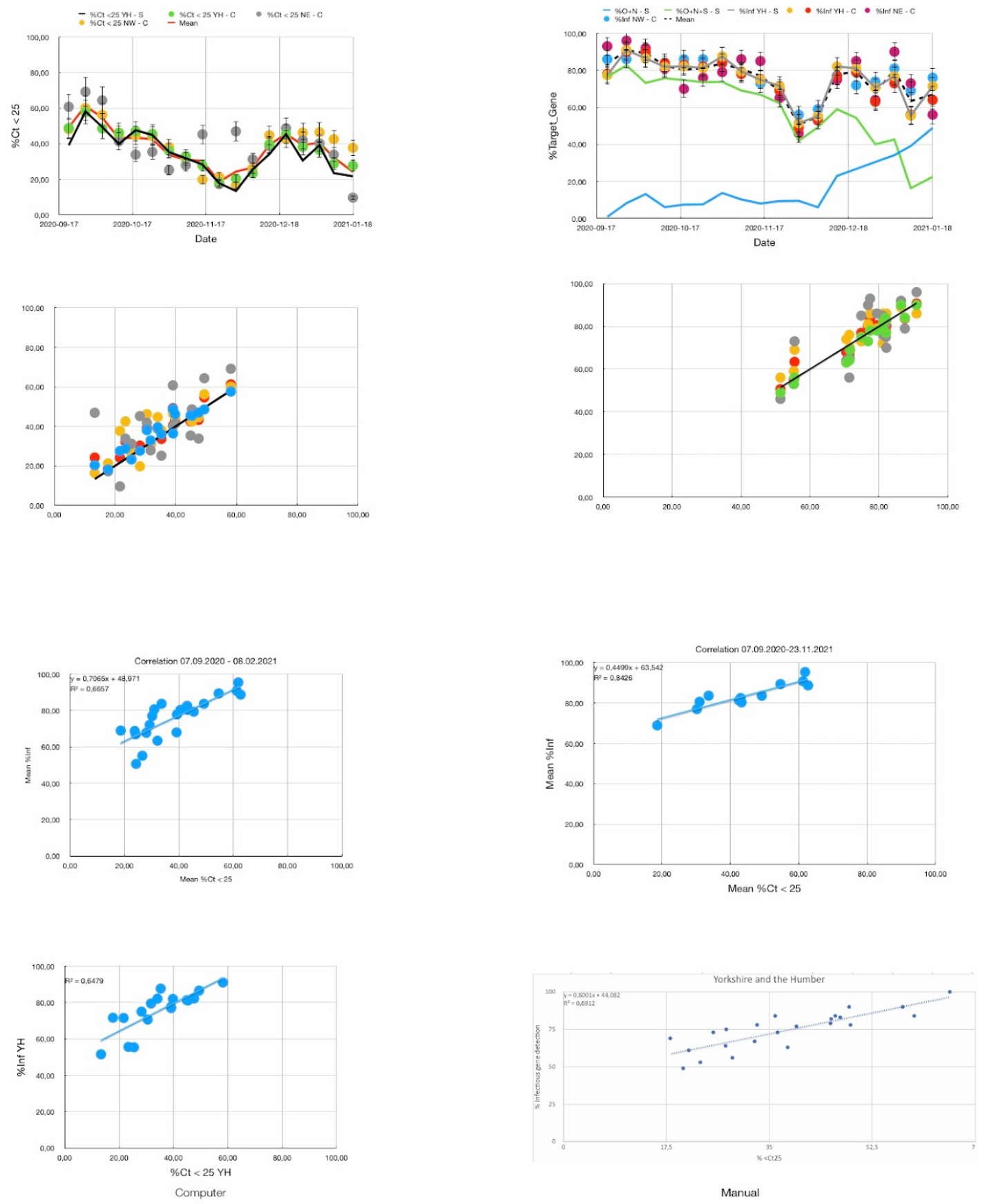
%Ct < 25 and %Inf: North

For the North, the computer generated %Ct < 25 for YH (Computer) was used as a reference against which the manually generated data for YH, NE, NW could be compared. This is shown in top left graph in the panel, where the black line represents the computer generated data and the points with error bars the manual data. The error bars correspond to an inter-quartile range of 3 cycles at Ct ∼ 25 giving a percentage error of 12%. Given the good match between the sets of data, it seems legitimate to calculate the mean for these sets, and this is shown as the red line in the graph.

The top right graph plots computed %Inf as the black line in the graph along with %ORF1ab+N (green) and %ORF1ab+N+S (blue). The points with error bars are the results obtained manually using Excel for YH, NE, NW. The error bars were estimated by eye as about 5% using the difference between the black line and the points on average. Again, the match was good between the sets of data, and the mean is plotted as the dashed black line.

The second row of graphs gives the correlation between the manual (y-axis) and computed data (x-axis) for both %Ct < 25 and %Inf. In the third row, the left graph gives the correlation between mean %Inf and mean Ct < 25 for the whole period of study 07.09.2020 - 08.02.2021. The co-efficient of determination, R^2^ = 0,666. However, this rises to 0,843 for the period before 23.11.2020 as demonstrated in the second graph on the right. The bottom row of graphs gives the correlations for YH as calculated by computer (left) and manually (right).

The analysis was repeated for the Midlands using the computer generated %Ct < 25 for WM as the reference line. Again, good matches were obtained and means calculated. In contrast to the North, there was no improvement obtained in R^2^ by restricting the time period. So the third row only contains one graph. For LSE, London was used as the reference. There was marginal improvement in R^2^ by restricting to the period after 16.11.2020.

The results can be summarised with the following plots:

**Figure 3:**
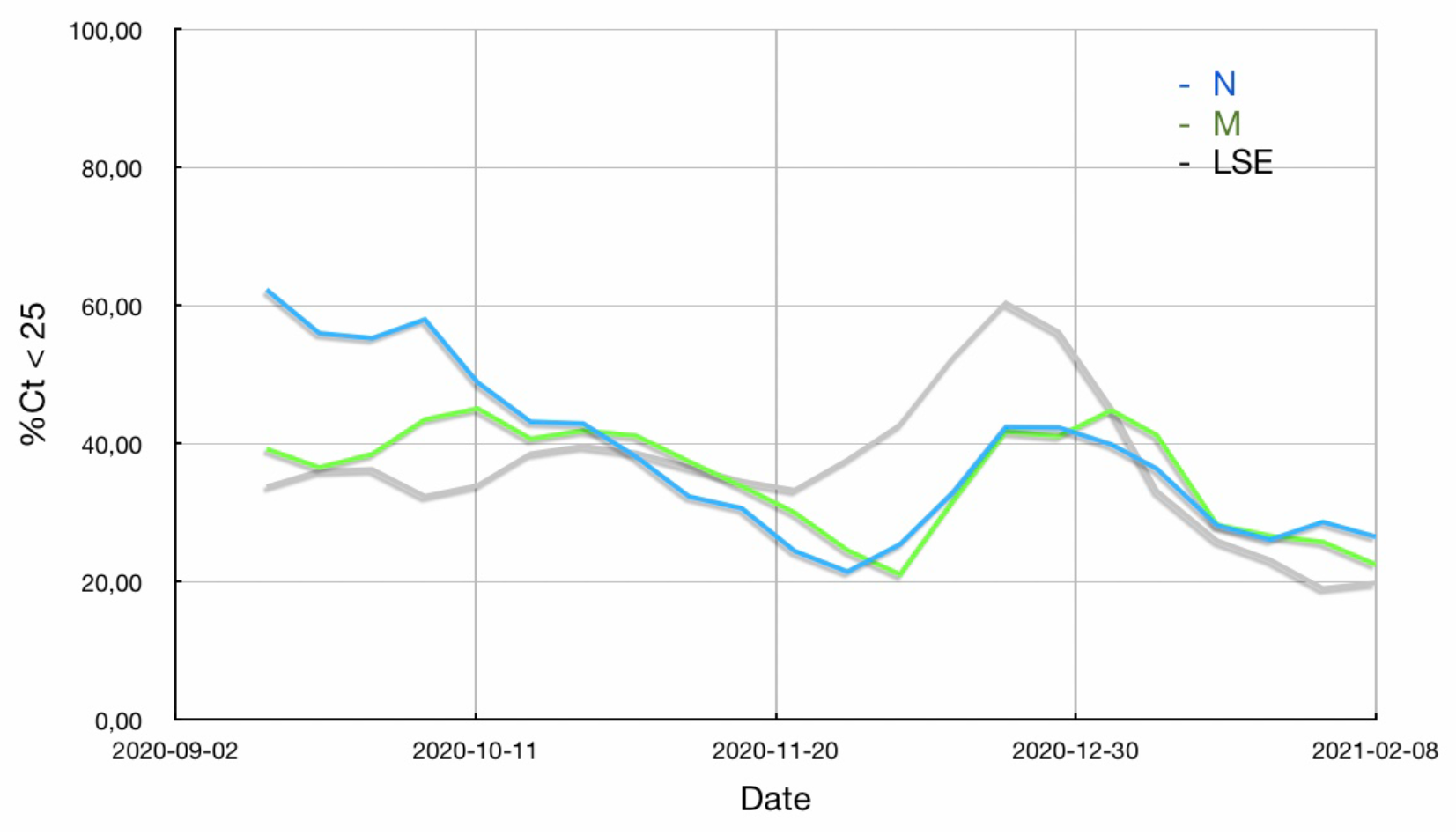
Mean %Ct < 25 v Date for the North (N), Midlands (M), London, South East and East England (LSE)

**Figure 4:**
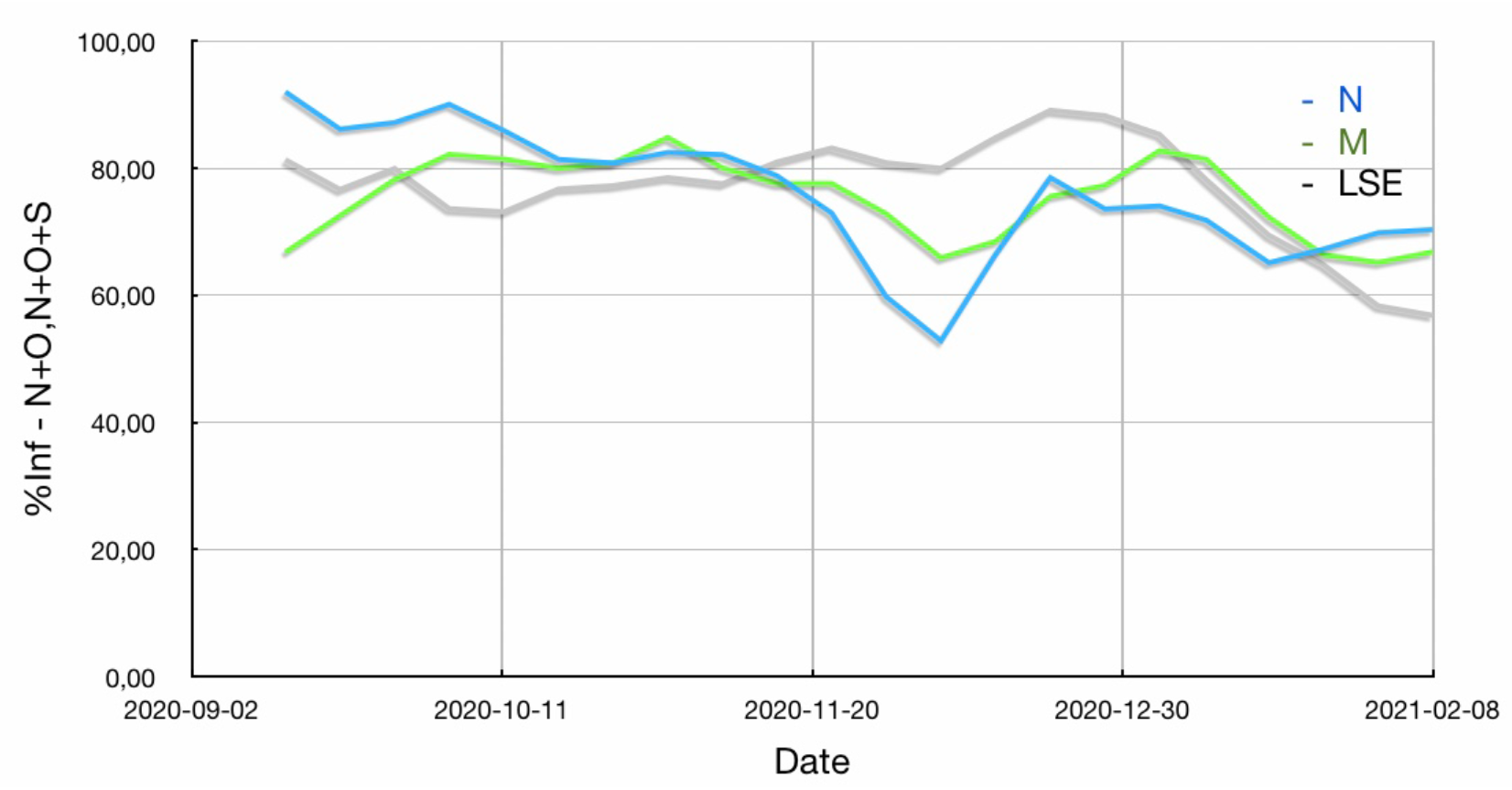
Mean %Inf v Date for the North (N), Midlands (M), London, South East and East England (LSE)

**Figure 5:**
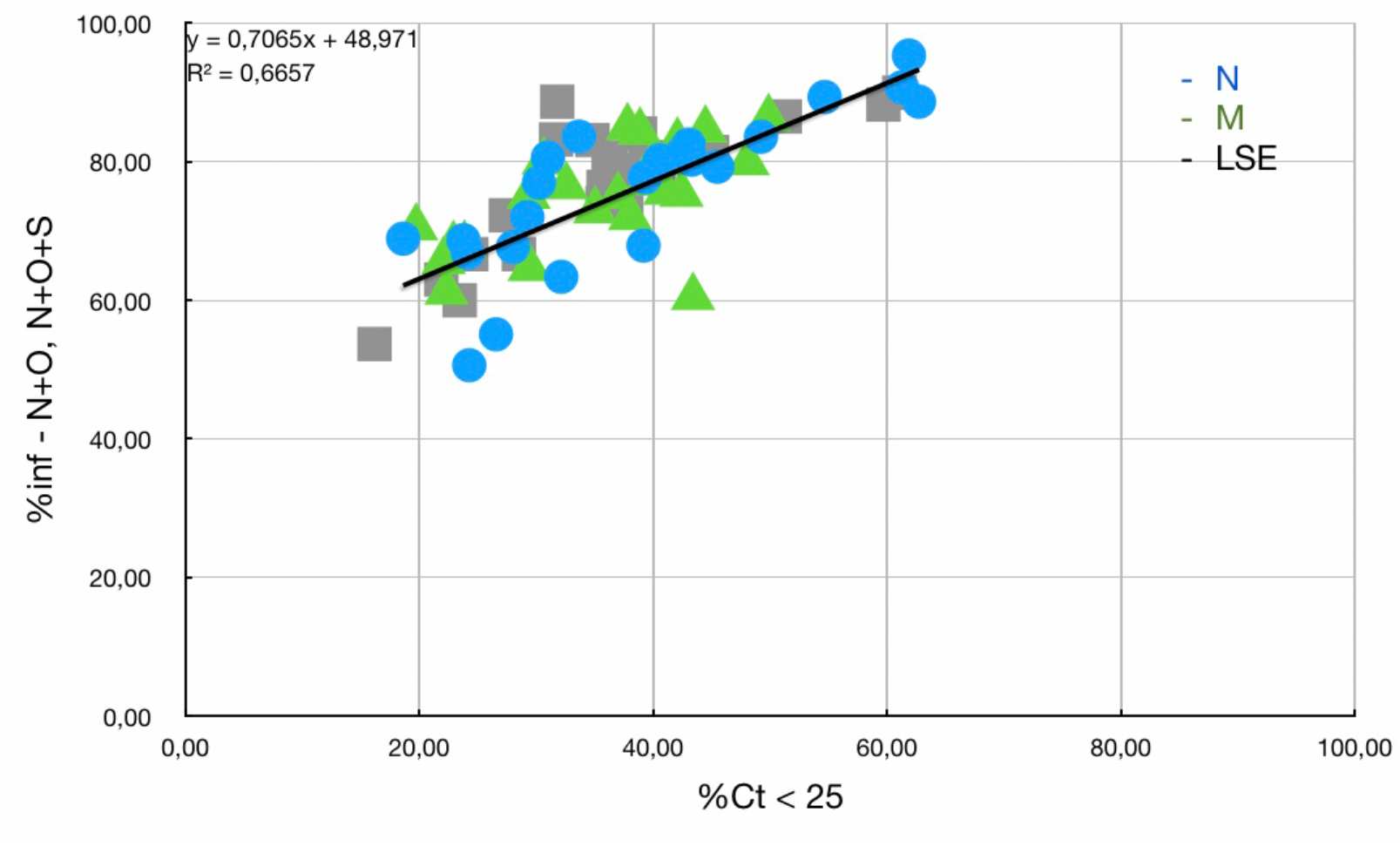
Cross-correlation: Mean %Inf v Mean %Ct < 25 for the North (N), Midlands (M), London, South East and East England (LSE)

Looking at the first graph, Mean %Ct < 25 v Date, we can see how the hot spot in the summer in the North moved southwards petering out at the end of November. This was followed by the rise of alpha in LSE in December, spreading to the other regions in January. The relatively strong correlation between %Ct<25 and %Inf confirms the biological basis for studying the former parameter.

## III. P(Ct) Distributions

The focus so far has been on the percentage of tests with Ct < 25, %Ct < 25, which was derived from plotting the ogive or cumulative frequency curve from the weekly ONS data. But the ogive just represents the integral of the underlying probability distribution, P(Ct), the probability of a positive test with a score Ct [8]. Further insight can be gained by studying these distributions.

The underlying distribution can be recovered simply by differentiating the ogive. By way of example, the ogive for London on 21.09.2020 is given below:

**Figure 6:**
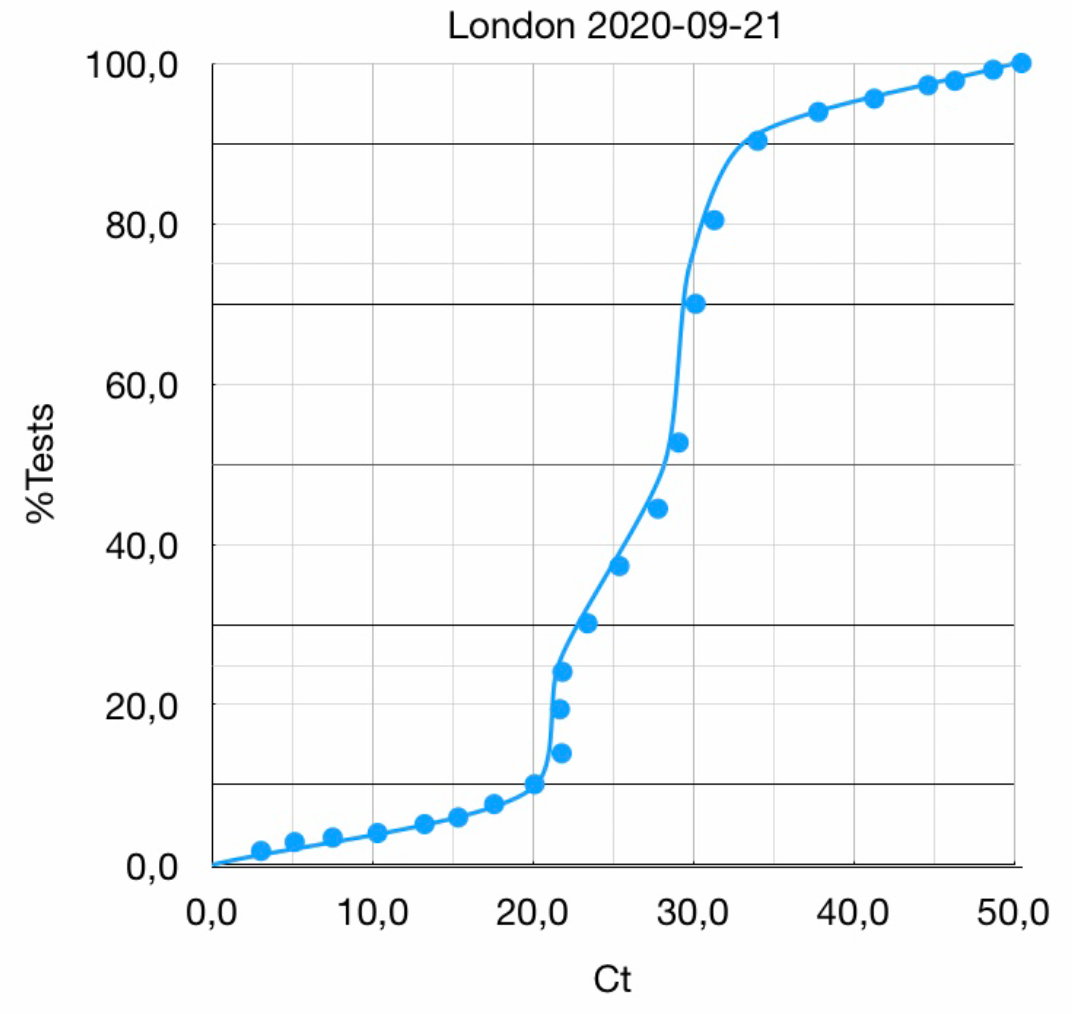
%Tests v Ct CMF for London 2020-09-21

which, after numerical differentiation using a standard five point difference formula, yields the following distribution:

**Figure 7:**
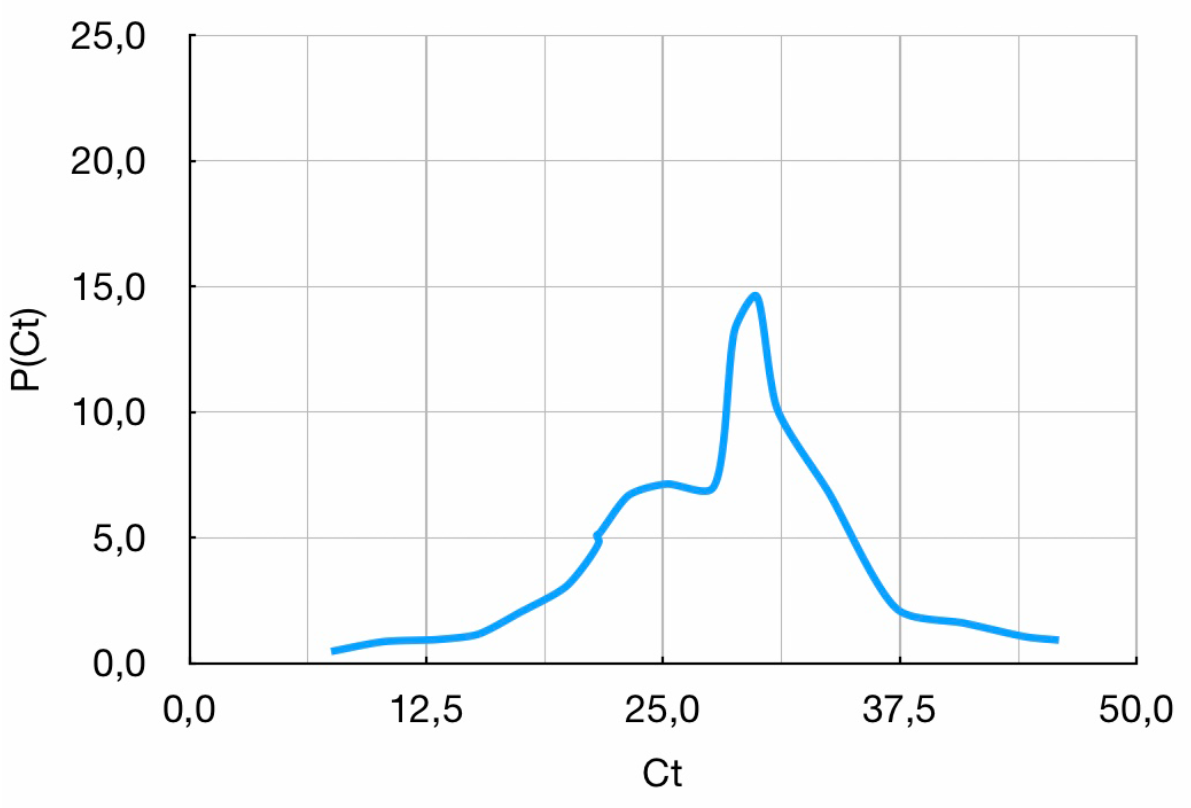
Probability Distribution P(Ct) v Threshold Score Ct for London 2020-09-21

The process can be automated using the R statistical programming language [22] with Tidyverse [23] and ggridges [24] packages to give the following results for the English regions between 21.09.2020 and 18.01.2021:

**Figure 8:**
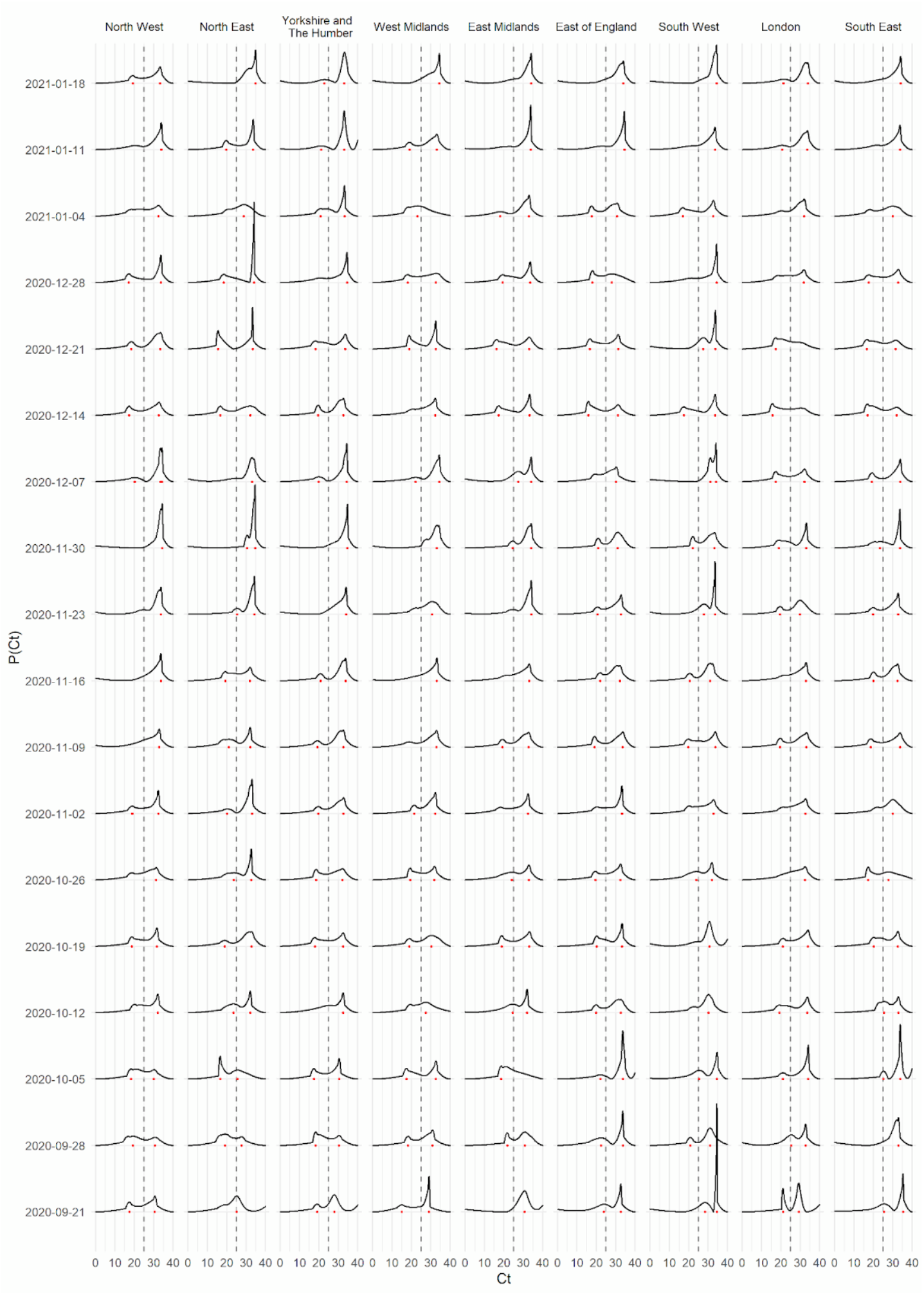
ONS P(Ct) Distributions for the English Regions v Date

The figure shows a ridgelines plot of P(Ct) distributions from the ONS community infection survey for the English Regions. Each line shows the distribution of Ct values on the date shown on the left-hand side. Each column shows the English region. The horizontal axis on each sub-plot shows the Ct value. The vertical axis of each sub-plot shows the relative proportion of each Ct value. The vertical axis scales have been removed for clarity, but the values of each sub-plot range between 0 and 1 and are scaled identically. Dots on the baseline of each sub-plot show the positions of the main peaks identified in each distribution. The data source is https://www.ons.gov.uk/peoplepopulationandcommunity/healthandsocialcare/conditionsanddiseases/bulletins/coronaviruscovid19infectionsurveypilot/29january2021

What immediately strikes the eye of the reader is the predominantly <u>bimodal</u> nature of the distributions.

Also published by ONS over a restricted range of time are percentiles for specific gene targets ORF1ab, N and S and their combinations. These again can be used to construct ogives which can be differentiated in a similar manner. The results are shown in the following plot:

**Figure 9:**
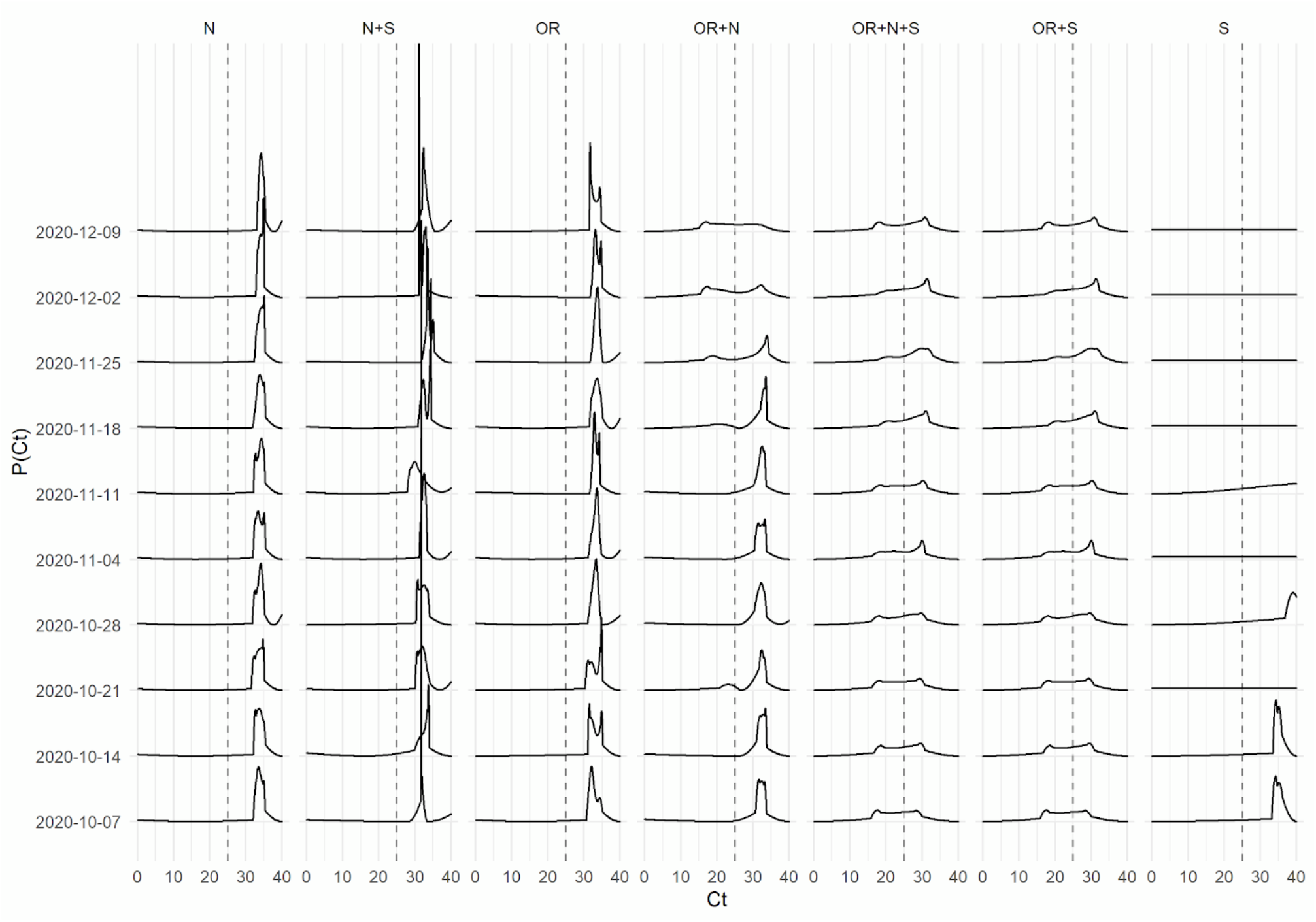
ONS P(Ct) Distributions for Gene Targets and Combinations v Date

The figure shows a ridgeline plot of ONS P(Ct) distributions for gene targets and combinations by date. Each line of sub-plots shows the distribution of each possible combination of positive PCR target identified each week for England. Each column of sub-plots shows the PCR target combinations (N, N+S, ORF1ab [OR], OR+N, OR+N+S and S). Note that due to an error in the dataset the data for OR+N+S and OR+S are identical. The horizontal axis on each sub-plot shows the Ct value. The vertical axis of each sub-plot shows the relative proportion of each Ct value. The vertical axis scales have been removed for clarity, but the values of each sub-plot range between 0 and 1 and are scaled identically.The data source is https://www.ons.gov.uk/file?uri=/peoplepopulationandcommunity/healthandsocialcare/conditionsanddiseases/adhocs/12692covid19infectionsurveyctanalysis/adhocctvalues.xlsx

Once more, the combinations ORF1ab+N, ORF1ab+N+S and ORF1ab+S, indicative of a positive result according to the established protocol, show predominantly a bimodal structure.

To shed light on the situation, it is instructive to plot the peak positions in the distributions against date. This is done for the regions N= NE, NW, YH, M = WM, EM, SW and LSE = L, SE, EE and gene targets ORF1ab+N, ORF1ab+N+S in Annex 1. The results for LSE are shown below:

**Figure 10:**
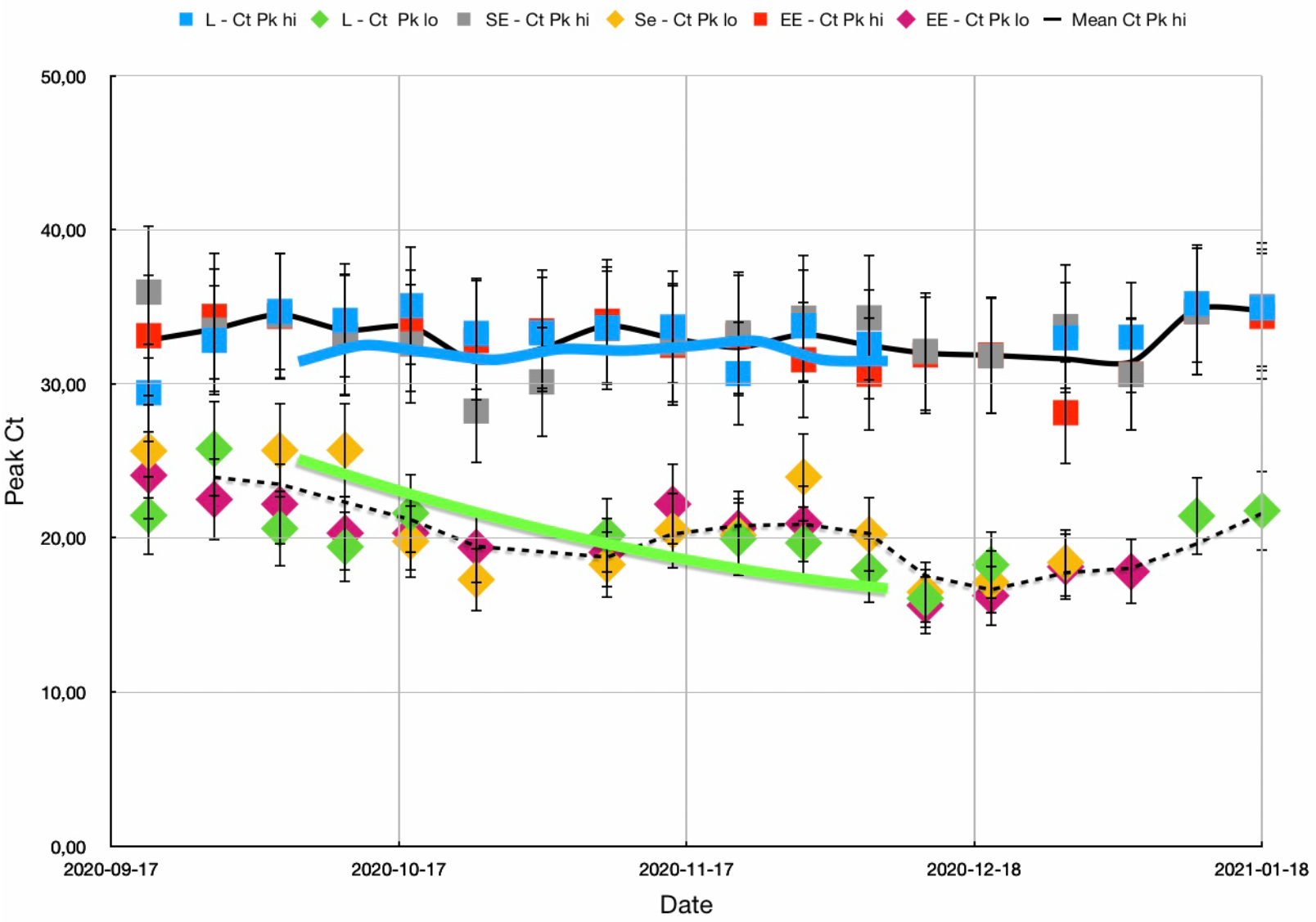
Peak Ct v Date for the English Regions London (L), South East (SE) and East England (EE)

The graph shows two series, one at a higher value of Ct (∼32) and one at a lower value of Ct (∼20). The black full line represents the mean of the three high values for London, L-Ct Pk hi, South East, SE-Ct Pk hi and East England, EE-Ct Pk hi at each date. The dashed line shows the mean of the lower values, L-Ct Pk lo, SE-Ct Pk lo and EE-Ct lo. The higher series remains essentially constant over time, with an average value of Ct = 32.95+/-1.11. The lower series apparently decreases with time, with the onset of the alpha variant, before increasing into the new year. The average value is Ct = 20.31+/-4.65. The large value for the standard deviation reflects the decreasing slope. Almost identical results are obtained for the North and the Midlands, with average values 32.24, 20.44 and 32.44, 20.12 respectively. The error bars correspond to an interquartile range of ∼ 3 cycles or 12%.

For comparison, the thick blue and green lines show the high and low peak values obtained for ORF1ab+N target genes, ORF1ab+N Ct Pk hi and ORF1ab+N Ct Pk lo respectively. These are shown in more detail in the following graph:

**Figure 11:**
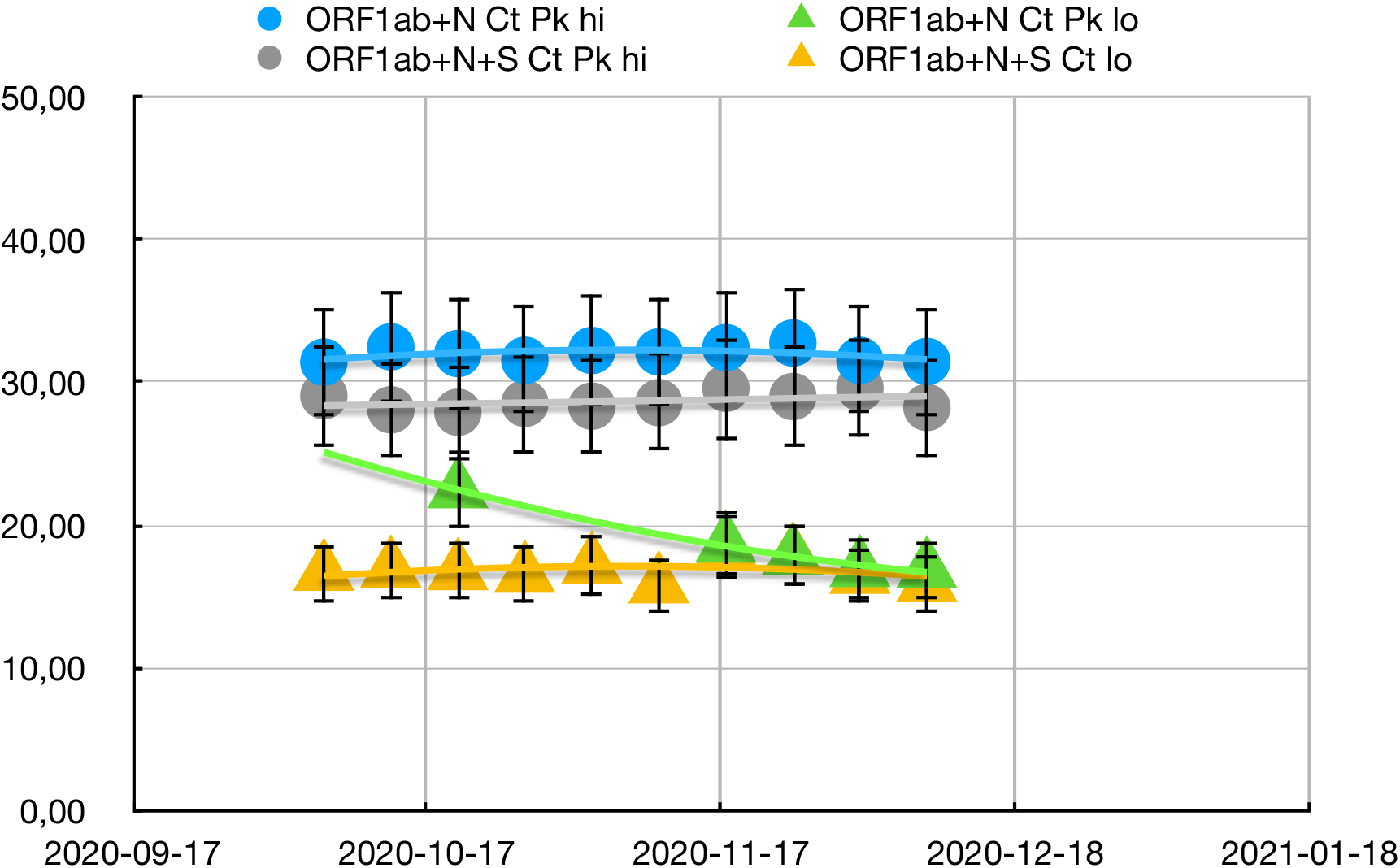
Peak Ct v Date for Gene Targets

The average value for the high series is 32.02+/-0.49 and that for the low series is 18.54+/-2.31. The black and yellow lines give very similar results for ORF1ab+N+S. The thick green trend line confirms the decrease in the lower series seen in the regional graphs.

Thus the P(Ct) distributions are characterised by a bimodal structure with a hot peak at a Ct value ∼ 20 and a cold peak with Ct ∼ 32, which closely match the peaks found for the target genes ORF1ab+N. Presumably, the cold peak corresponds to residue left over from previous infections, whereas the hot peak is indicative of a current infection. This is discussed below.

## IV. Discussion

The presence of two peaks in the Ct distributions requires some discussion. The fact that the distributions are reported as percentiles rather as the underlying distributions suggests that only unimodal/normal distributions have been contemplated by software designers. Indeed the distributions for the single gene targets are unimodal (cf. figure 7 above). So what is the source of the two peaks?

The assumption in the literature [10,11] seems to be that before a person develops symptoms, a low viral load will be detectable with a high Ct score, > 30 say, followed by a low score, < 25, as the viral load increases, and then a high score again post-infection. This indicated in the figure below taken from reference [10].

**Figure 12:**
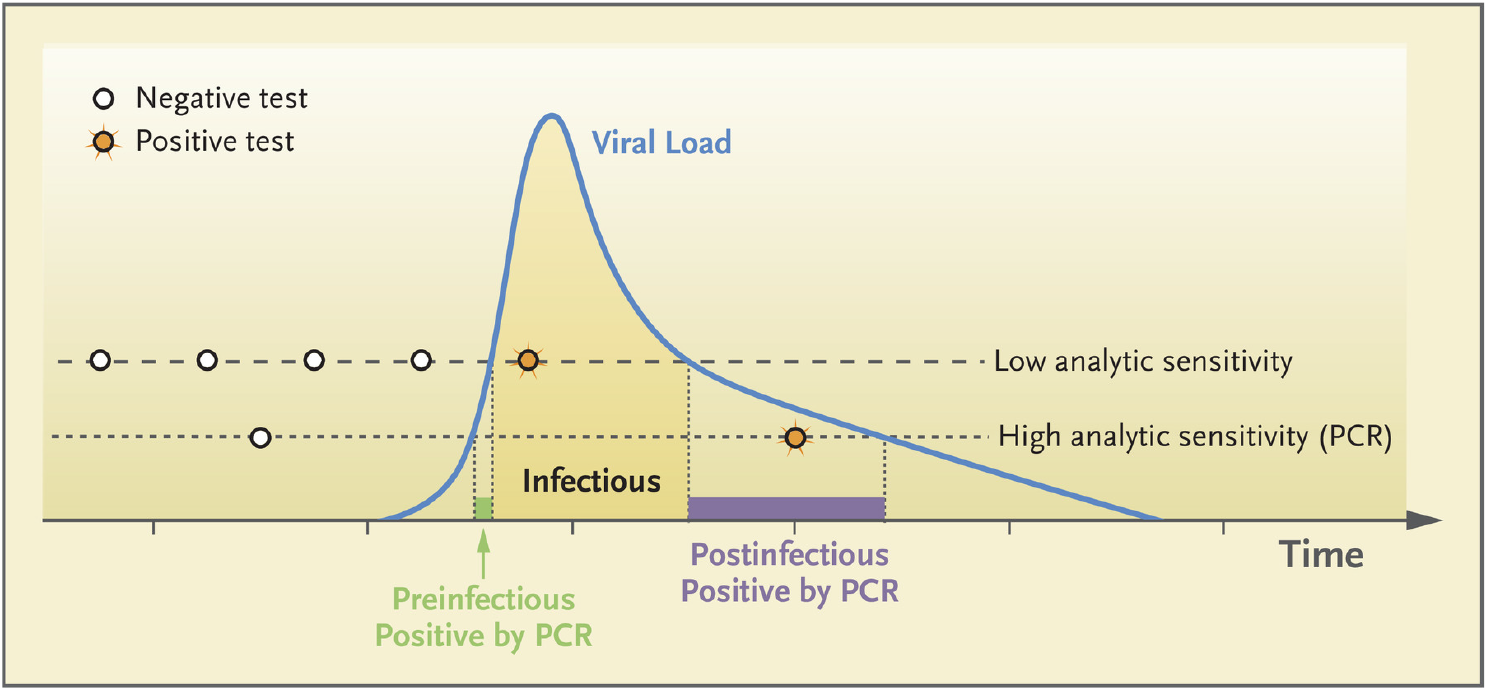
Trajectory of Viral Load v Time and its Effects on Test Results

However, in our view, this signal when convoluted with the distribution of people under test would not give a bimodal distribution but rather a curve with a single peak and a long tail to high Ct.

In any case, the distributions for the single gene targets show a single peak at Ct > 30, and the peak at Ct ∼ 20 only occurs in the distributions for a combination of targets, OR+N, OR+N+S, the criterion for a positive test. Thus it seems fair to infer that only the peak at Ct ∼ 20 can be associated with live viral load and the peak at ∼ 32 would seem to represent residue left over from a previous infection.

The ONS data for specific gene targets and their combinations have also been analysed by Walker et al [12]. The aggregate distributions for the time period 26.04.2020 - 13.03.2021 are displayed below in the form of violin plots:

**Figure 13:**
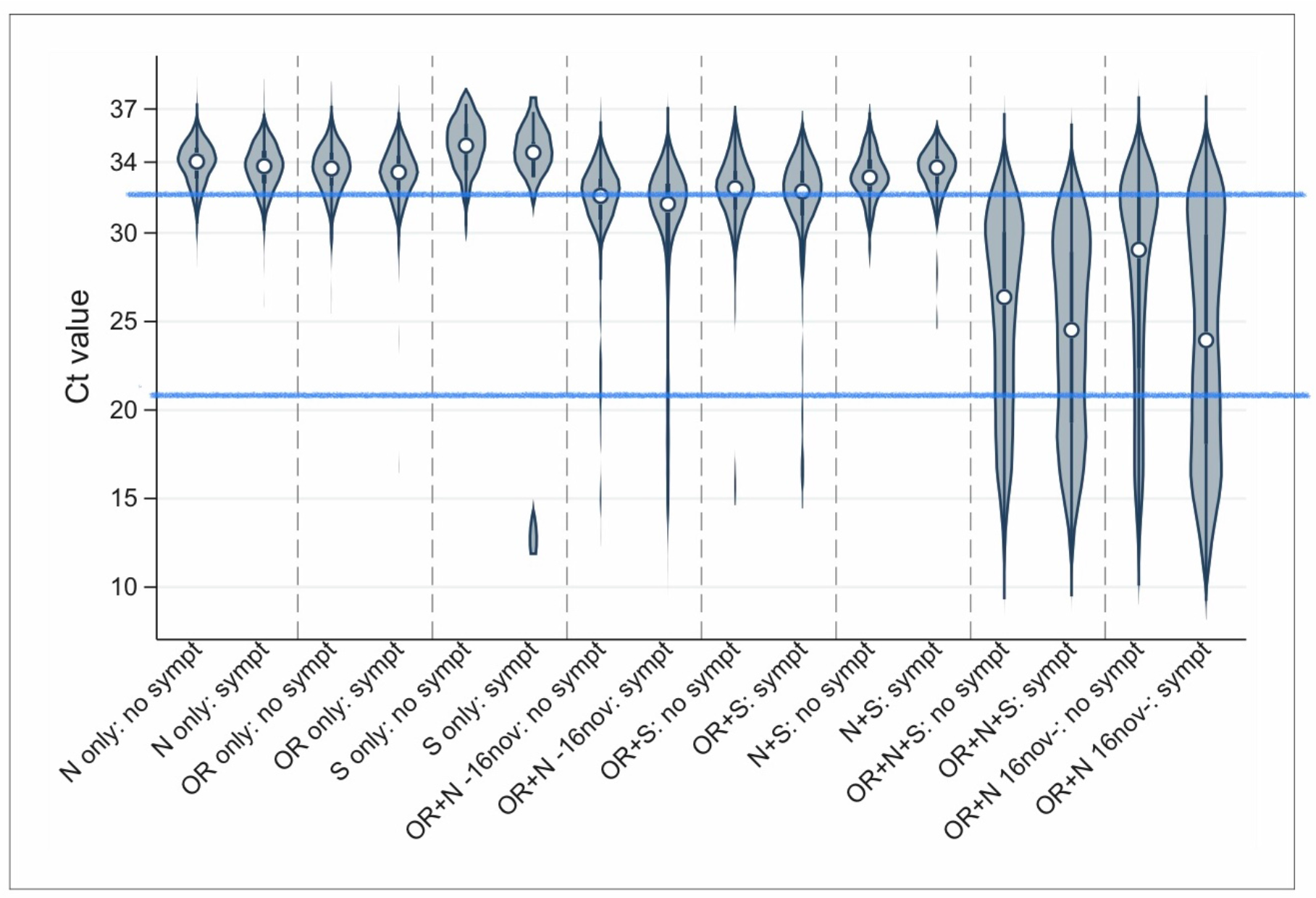
Distributions of Ct values by genes detected

For comparison, the blue lines represent the mean values for the high and low series given above. In confirmation of the present work, the single gene targets in [12] also show a single peak at Ct > 30 whereas the combination targets OR+N, OR+N+S etc. are clearly bimodal with a second peak at Ct < 20. According to the authors in [12] lower Ct values likely reflect the natural history of viral load post-infection, whereas higher Ct values in those testing positive indicate long-term shedders.

In any case, there have been other reports of bimodal distributions in references [13,14]. In [13], the two peaks for N1 and N2 genes were quantitatively measured by fitting a mixture of two normal distributions. This yielded a mean of ∼ 20 for the first peak and ∼ 33 for the second. Reference [14] found that distributions of OR and N Ct values in screening cases were bimodal. It was hypothesised that the lower peak corresponded to patients with high infectivity whereas the higher peak was due to patients with low infectivity. The distributions were modelled using a maximum likelihood method assuming a mixture of Gaussians in order to establish an effective Ct cutoff value. For the ORF1ab gene, the population means were 25.3 and 33.8; for the N gene, 26.0 and 34.9.

We have also determined the distributions for various gene targets and combinations from the cfrvoc Pillar 2 dataset [15,16]:

**Figure 14:**
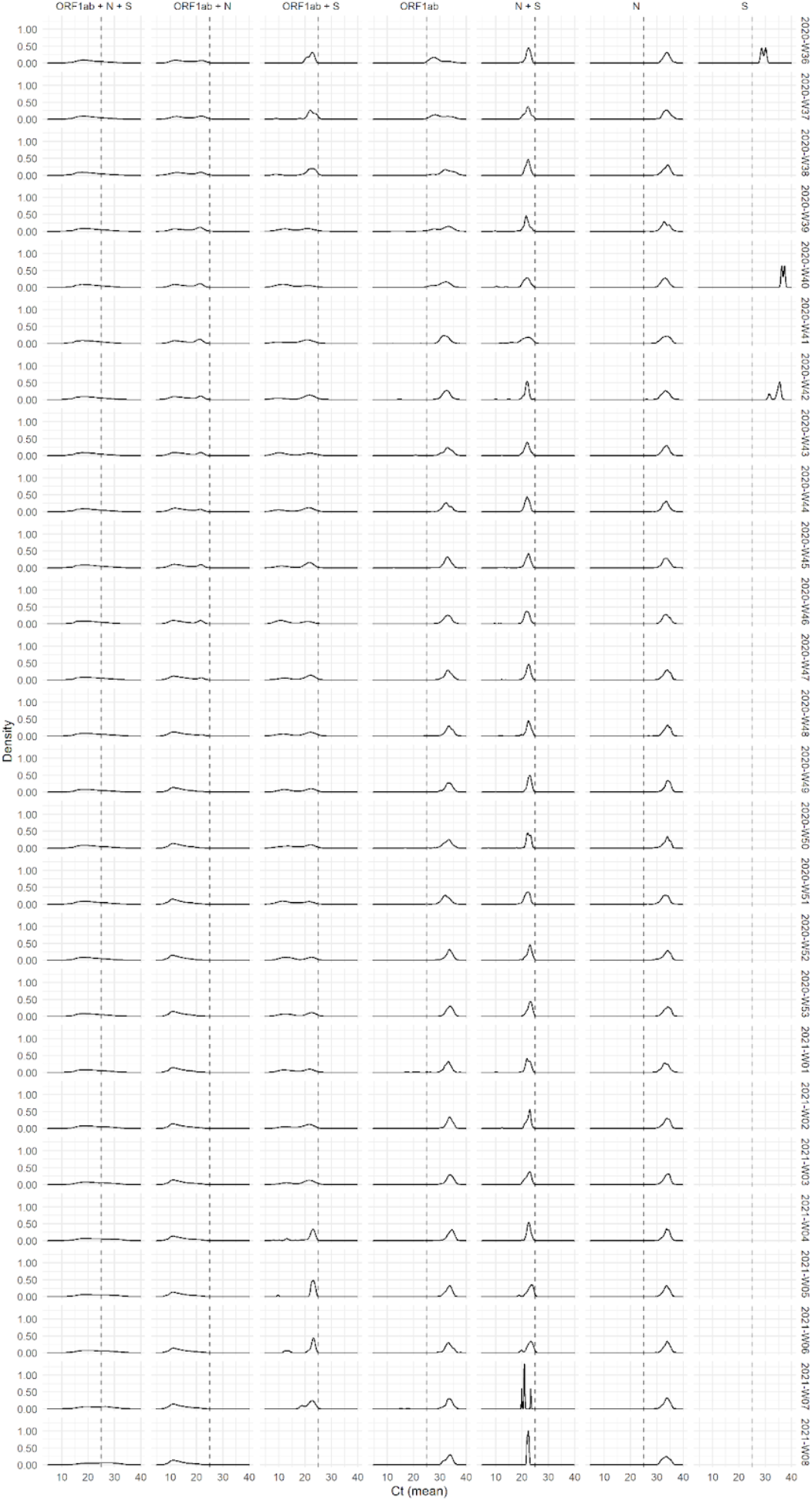
cfroc Pillar 2 P(Ct) Distributions for Gene Targets and Combinations v Date

Again the distributions for ORF1ab+N, ORF1ab+N+S are predominantly bimodal but with peaks at substantially lower values of Ct (< 25) whereas those for the single genes remain unimodal. As the ONS and Pillar 2 use the same laboratories, the difference in peak values can only be ascribed to the differences in the population sampled. ONS samples the general population while Pillar 2 is supposed to reflect people with symptoms. So it is perhaps not surprising that peaks occur at lower values in the CFRVOC results.

Thus, as suggested in references [13,15], the bimodal Ct distribution can be used in test reports as a relatively simple indicator that can be useful for the management of infected patients. Clearly, however, the exact values remain apparatus and sample dependent. This emphasises the need for calibration.

## V. Conclusions

We have demonstrated that a significant parameter for tracking the course of COVID in the second wave is the percentage of positive tests with Ct < 25. The biological basis for studying this parameter is the strong correlation between %Ct < 25 and the percentage of positive tests comprising target genes ORF1ab+N and ORF1ab+N+S, or %Inf. Indeed, %Ct < 25 can be combined at national and regional levels with pillar 2 results to estimate the cumulative total of cold positives, ie. non-infectious testing positive, in the second wave [17], which concurs with the estimate from a scaling analysis [18].

Furthermore, the probability distributions, giving the probability of a positive test with a score Ct, P(Ct) were found to be predominantly bimodal with a hot peak at a Ct value ∼ 20 and a cold peak with Ct ∼ 32, which closely match the peaks found for the target genes ORF1ab+N. Similar results were found in [13] and [14]. As discussed above, the cold peak cannot be assigned to low viral load of a current infection, which later becomes infectious. Rather it seems more likely associated with residue from a previous infection. The distributions for gene targets in CFRVOC are also bimodal but the peaks occur at lower values of Ct. This suggests the results are machine/sample dependent and emphasises the need for calibration, if quality control in PCR testing is to be improved. See also the latest recommendations of Heneghan et al in reference [19].

## Supporting information

Ct vs Inf datasets and graphs

P(Ct) datasets and graphs

## Data Availability

All data produced in the present study are available upon reasonable request to the authors

https://drive.google.com/file/d/1HlRrd5b-gcSib1362brJ92kf6ch-kLoQ/view?usp=drivesdk

https://drive.google.com/file/d/1vkptBSjjUstTr2ZIddk0IyAJdrs1v1fH/view?usp=drivesdk

## Acknowledgements

We are grateful for the preliminary data preparation by SHU students Alice Atkinson, Rachel Davies and Charlotte Simmonds performed as part of their final year dissertations.

## Annex I. Methods and Data Analysis

Data used in this study were obtained from the official databases from the Office of National Statistics (ONS) COVID Infection survey (CIS) between the 01.09.2020 and 12.02.2021, which correlated with the second peak of COVID-19 infections in the UK (cf. https://www.ons.gov.uk/peoplepopulationandcommunity/healthandsocialcare/conditionsanddiseases/datasets/coronaviruscovid19infectionsurveydata?s=03). Data were extracted for all 9 regions across England: North East, North West, Yorkshire & Humber, West Midlands, East Midlands, East of England, London, South East and South West, downloaded and stored in Excel spreadsheets by our students (AA, RD, CS) for preliminary analysis.

In particular, weekly Ct percentile values were collected from table 6B of the ONS CIS over a five-month period which began the week commencing 07.09.2020 and finished the week commencing 08.02.2021. The weekly Ct percentiles (y axis) were plotted against the Ct values (x axis) to create the cumulative frequency function (CMF) or ogive (cf. fig 1 above). The curve was fitted with a cubic polynomial to yield %Ct <25 values. Inconsistencies were resolved manually, e.g. by linear interpolation.

The different gene detection percentages were collected over the same five-month period. Positive tests for combined ORF1AB + N + S genes or ORF1AB + N genes were deemed to be infectious; other positive tests that detected only N, only S, only ORF1AB proteins or N + S or ORF1AB + S proteins were deemed non-infectious especially since positives based on S gene detection are now held to be unreliable, following WHO guidance [20,21]. The percentage of positive tests assigned to infectious gene combinations, %Inf was calculated by dividing the number of infectious tests by the sum of the non-infectious and infectious positive values. A preliminary comparison of %Ct with %Inf showed the data for 07.09.2020, 14.09.2020, and after 18.01.2021 constituted outliers and were discarded [17].

In parallel, the same data were downloaded and analysed using R statistical programming language [22]. Again ogives were constructed and fitted with cubic splines to yield %Ct < 25 values. The curves were then differentiated numerically to give the underlying P(Ct) distributions (cf. fig.8 above). By fitting Gaussians to the individual peaks of the distributions the area under each peak could be calculated and the peak maxima determined.

In similar fashion, ogives for the infectious gene combinations were determined and the curves differentiated to yield the underlying P(Ct) distributions and peak maxima were determined (cf. fig. 9). The percentages of positive tests with targets ORFlab + N + S and ORFlab + N were also calculated. The cfroc data were treated analogously (cf. fig. 14).

The various sets of data for %Ct < 25, %Inf, peak maxima and area were copied to Numbers spreadsheet *SHU Collaboration* and analysed to yield the results discussed above. The spreadsheet is available on iCloud upon reasonable request to the authors.

## Annex II. Infectious Genes

https://drive.google.com/file/d/1HlRrd5b-gcSib1362brJ92kf6ch-kLoQ/view?usp=drivesdk

## Annex III. P(Ct) Distributions

https://drive.google.com/file/d/1vkptBSjjUstTr2ZIddk0IyAJdrs1v1fH/view?usp=drivesdk

## References

[1] Office of National Statistics COVID-19 Infection Survey. https://www.ons.gov.uk/surveys/informationforhouseholdsandindividuals/householdandindividualsurveys/covid19infectionsurvey

[2] Yeadon M, https://lockdownsceptics.org/addressing-the-cv19-second-wave/

[3] Jefferson T, Spencer EA, Brassey J, Heneghan C, ‘Viral cultures for COVID-19 infectivity assessment - a systematic review’, medRxiv preprint, doi: https://doi.org/10.1101/2020.08.04.20167932

[4] https://www.gov.uk/government/publications/sage-73-minutes-coronavirus-covid-19-response-17-december-2020

[5] Bullard J, Dust K, Funk D, et al. ‘Predicting infectious severe acute respiratory syndrome corona-virus 2 from diagnostic samples’, Clin.Infect. Dis. 2020; doi:10.1093/cid/ciaa6

[6] https://www.corriere.it/cronache/20_giugno_19/coronavirus-remuzzi-nuovi-positivi-non-sono-contagiosi-stop-paura-bf24c59c-b199-11ea-842e-6a88f68d3e0a.shtml

[7] ‘Datasets: Coronavirus (COVID-19) Infection Survey: England (12 March 2021)’, Coronavirus (COVID-19) Infection Survey: England, Mar. 12, 2021. https://www.ons.gov.uk/file?uri=/peoplepopulationandcommunity/healthandsocialcare/conditionsanddiseases/datasets/coronaviruscovid19infectionsurveydata/2021/previous/v12/covid19infectionsurveydatasets20210305v2.xlsx

[8] See https://en.wikipedia.org/wiki/Ogive_(statistics); https://en.wikipedia.org/wiki/Cumulative_distribution_function

[9] SHU Collaboration reports: A Compendium, https://drive.google.com/file/d/1oaJ3j7V32340VnLnJaPVWUEBUcq3IsRS/view?usp=drivesdk

[10] Mina MJ et al, N Engl J Med 2020; 383:e120, DOI: 10.1056/NEJMp2025631

[11] LADR Laborverbund Dr Kramer & Kollegen, ‘SARS-Coronavirus-2 PCR: Wie aussagekräftig ist der Ct-Wert?’, https://ladr.de/ct-wert-sars-coronavirus-2-pcr

[12] Walker AS, Pritchard E, House T, Robotham JV, Birrell PJ, Bell I, et al. Ct threshold values, a proxy for viral load in community SARS-CoV-2 cases, demonstrate wide variation across populations and over time. eLife. 2021 Jul 12;10:e64683

[13] Yang D et al ‘Bimodal Distribution Pattern Associated With The PCR Cycle Threshold (Ct) And Implications In COVID-19 Infections’, https://doi.org/10.21203/rs.3.rs-1340499/v1

[14] Khelil Mohamed Mokhtar, ‘Improved RT-PCR SARS-Cov2 results interpretation by indirect determination of cut-off cycle threshold value’, medRxiv preprint, https://doi.org/10.1101/2020.11.20.20235390

[15] nicholasdavies. nicholasdavies/cfrvoc. 2021. Available from: https://github.com/nicholasdavies/cfrvoc

[16] Davies N. CSV-format data for: Increased mortality in community-tested cases of SARS-CoV-2 lineage B.1.1.7. Zenodo; 2021. Available from: https://zenodo.org/record/5105921

[17] Simmonds C, Final Year Dissertation,Sheffield Hallam University, May, 2021; Johnson K, https://drive.google.com/file/d/1q4xlaJ1V4c67JHIupwM7yf1HMVhYUQ9u/view?usp=drivesdk.

[18] Johnson K, ‘A Scaling Law for PCR Positivity in the COVID Second Wave’, medRxiv preprint, https://doi.org/10.1101/2021.12.07.21267073; Advance Research Journal of medical and clinical science. 2022;802–810, https://doi.org/10.15520/arjmcs.v8i04.426

[19] Jefferson T, Dietrich M, Brassey J, Heneghan C, ‘PCR Testing in the UK During the SARS-CoV-2 Pandemic – Evidence from FOI Requests’, medRxiv preprint, https://doi.org/10.1101/2022.04.28.22274341

[20] Pouwels, K. B., House, T., Pritchard, E., Robotham, J. V., Birrell, P. J., Gelman, A., Kerby, M., 2021, ‘Community prevalence of SARS-CoV-2 in England from April to November, 2020: Results from the ONS coronavirus infection survey. The Lancet. Public Health, 6(1), e30–e38. doi:10.1016/S2468-2667(20)30282-6.

[21] The World Health Organisation, 2020 ‘WHO Emergency Use Assessment Coronavirus Disease (COVID-19) IVDS Public Report’: https://www.who.int/diagnostics_laboratory/eual200921_final_pqpr_eul_0525_156_00_taqpath_covid19_ce_ivd_rt_pcr_kit.pdf?ua=1

[22] R Core Team (2021). ‘R: A language and environment for statistical computing’. R Foundation for Statistical Computing, Vienna, Austria. URL https://www.R-project.org/.

[23] Wickham et al., (2019), ‘Welcome to the tidyverse’, Journal of Open Source Software, 4(43), 1686, https://doi.org/10.21105/joss.01686.

[24] Claus O. Wilke (2021), ‘ggridges: Ridgeline Plots in ‘ggplot2’, R package version 0.5.3. https://CRAN.R-project.org/package=ggridges

