## Supplementary material for "An Analysis of PCR Ct Scores and Distributions from the ONS Community Infection Survey during the COVID Second Wave in the UK": Ct vs Inf datasets and graphs

### Summary

Table 1

| | Date | Mean %Ct <25 N | Mean %Ct < 25 M | Mean %Ct < 25 LSE | | Mean %Inf N | Mean %Inf - M | Mean %Inf - LSE |
|---|---|---|---|---|---|---|---|---|
| | 2020-09-07 | 61,86 | 43,39 | 31,78 | | 95,33 | 60,50 | 88,67 |
| | 2020-09-14 | 62,73 | 35,03 | 35,66 | | 88,67 | 73,00 | 74,00 |
| | 2020-09-21 | 49,21 | 38,02 | 36,34 | | 83,61 | 72,03 | 79,31 |
| | 2020-09-28 | 61,27 | 38,86 | 36,17 | | 90,73 | 84,36 | 80,61 |
| | 2020-10-05 | 54,66 | 48,07 | 28,51 | | 89,35 | 79,95 | 66,65 |
| | 2020-10-12 | 43,02 | 42,06 | 39,32 | | 82,47 | 82,93 | 79,63 |
| | 2020-10-19 | 43,27 | 39,31 | 37,69 | | 80,29 | 77,02 | 73,81 |
| | 2020-10-26 | 42,50 | 44,46 | 41,54 | | 81,31 | 84,66 | 80,67 |
| | 2020-11-02 | 33,64 | 37,79 | 35,74 | | 83,63 | 84,99 | 76,36 |
| | 2020-11-09 | 30,99 | 36,99 | 37,41 | | 80,60 | 75,08 | 78,74 |
| | 2020-11-16 | 30,22 | 30,63 | 31,68 | | 76,97 | 80,06 | 83,21 |
| | 2020-11-23 | 18,60 | 29,34 | 34,80 | | 68,89 | 75,10 | 83,12 |
| | 2020-11-30 | 24,26 | 19,72 | 40,42 | | 50,61 | 70,53 | 78,56 |
| | 2020-12-07 | 26,55 | 22,31 | 45,02 | | 55,08 | 61,26 | 81,39 |
| | 2020-12-14 | 39,28 | 41,03 | 59,68 | | 77,75 | 75,70 | 88,25 |
| | 2020-12-21 | 45,47 | 42,42 | 61,04 | | 79,27 | 75,39 | 89,97 |
| | 2020-12-28 | 39,16 | 39,73 | 51,25 | | 67,89 | 79,17 | 86,54 |
| | 2021-01-04 | 40,54 | 49,86 | 38,86 | | 80,21 | 86,31 | 84,07 |
| | 2021-01-10 | 32,12 | 32,57 | 27,39 | | 63,38 | 76,49 | 72,25 |
| | 2021-01-18 | 24,13 | 23,85 | 24,43 | | 66,85 | 68,08 | 66,64 |
| | 2021-01-25 | 27,99 | 29,42 | 21,86 | | 67,67 | 64,67 | 63,00 |
| | 2021-02-01 | 29,23 | 22,05 | 16,15 | | 72,00 | 65,67 | 53,67 |
| | 2021-02-08 | 23,76 | 22,91 | 23,44 | | 68,67 | 68,00 | 60,00 |

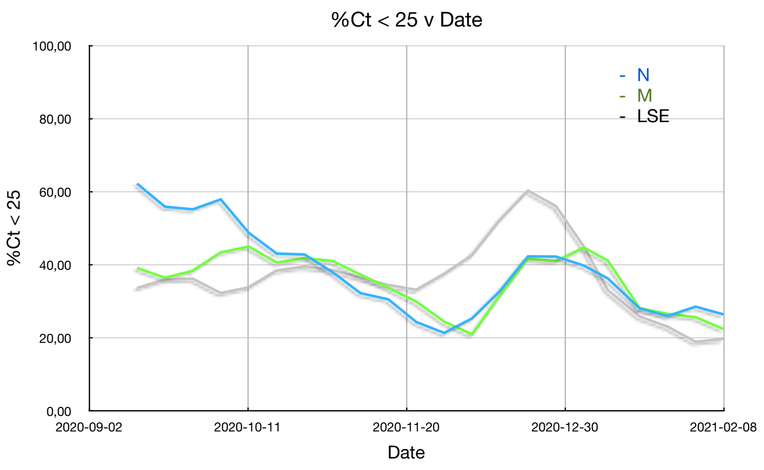

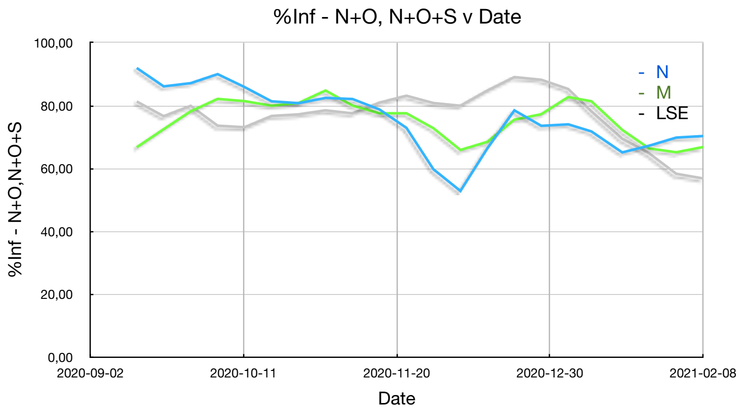
%Inf - N+O, N+O+S v Date
100,00
80,00
60,00
40,00
20,00
0,00
%Inf - N+O,N+O+S
2020-09-02
2020-10-11
2020-11-20
2020-12-30
2021-02-08
Date
- N
- M
- LSE

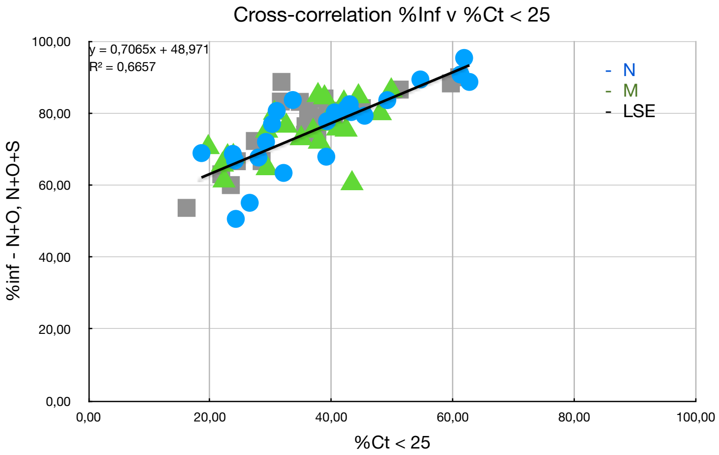
Cross-correlation %Inf v %Ct < 25
y = 0,7065x + 48,971
R² = 0,6657
100,00
80,00
60,00
40,00
20,00
0,00
%inf - N+O, N+O+S
0,00
20,00
40,00
60,00
80,00
100,00
%Ct < 25
- N
- M
- LSE

### North

Table 1

| Date | %Ct <25 YH - S | %Ct < 25 YH - C | %Ct < 25 NE - C | %Ct < 25 NW - C | Mean | %O+N - S | %O+N+S - S | %Inf YH - S | %Inf YH - C | %Inf NE - C | %Inf NW - C | Mean |
|---|---|---|---|---|---|---|---|---|---|---|---|---|
| 2020-09-07 | | 65,68 | 49,98 | 69,93 | 61,86 | | | | 100,00 | 100,00 | 86,00 | 95,33 |
| 2020-09-14 | | 59,63 | 74,88 | 53,67 | 62,73 | | | | 84,00 | 89,00 | 93,00 | 88,67 |
| 2020-09-21 | 39,08 | 48,80 | 60,77 | 48,18 | 49,21 | 0,91 | 76,52 | 77,44 | 78,00 | 93,00 | 86,00 | 83,61 |
| 2020-09-28 | 58,16 | 57,66 | 69,19 | 60,07 | 61,27 | 8,26 | 82,66 | 90,92 | 90,00 | 96,00 | 86,00 | 90,73 |
| 2020-10-05 | 49,41 | 48,58 | 64,37 | 56,26 | 54,66 | 13,20 | 73,20 | 86,40 | 90,00 | 92,00 | 89,00 | 89,35 |
| 2020-10-12 | 39,79 | 46,21 | 41,38 | 44,69 | 43,02 | 6,14 | 75,74 | 81,88 | 84,00 | 81,00 | 83,00 | 82,47 |
| 2020-10-19 | 47,56 | 47,05 | 33,83 | 44,65 | 43,27 | 7,47 | 74,68 | 82,16 | 83,00 | 70,00 | 86,00 | 80,29 |
| 2020-10-26 | 44,90 | 45,51 | 35,4 | 44,20 | 42,50 | 7,61 | 73,61 | 81,23 | 82,00 | 76,00 | 86,00 | 81,31 |
| 2020-11-02 | 35,28 | 35,99 | 25,17 | 38,11 | 33,64 | 13,76 | 73,76 | 87,52 | 84,00 | 79,00 | 84,00 | 83,63 |
| 2020-11-09 | 31,75 | 32,92 | 27,86 | 31,43 | 30,99 | 10,30 | 69,10 | 79,40 | 78,00 | 86,00 | 79,00 | 80,60 |
| 2020-11-16 | 28,22 | 27,67 | 45,23 | 19,78 | 30,22 | 8,04 | 66,84 | 74,88 | 75,00 | 85,00 | 73,00 | 76,97 |
| 2020-11-23 | 17,73 | 18,15 | 17,24 | 21,29 | 18,60 | 9,38 | 62,18 | 71,56 | 69,00 | 65,00 | 70,00 | 68,89 |
| 2020-11-30 | 13,33 | 20,35 | 46,92 | 16,43 | 24,26 | 9,51 | 41,93 | 51,44 | 49,00 | 46,00 | 56,00 | 50,61 |
| 2020-12-07 | 25,45 | 23,26 | 31,17 | 26,31 | 26,55 | 6,06 | 49,26 | 55,32 | 53,00 | 53,00 | 59,00 | 55,08 |
| 2020-12-14 | 34,09 | 39,60 | 38,75 | 44,70 | 39,28 | 23,01 | 59,01 | 82,02 | 77,00 | 75,00 | 77,00 | 77,75 |
| 2020-12-21 | 45,34 | 45,40 | 48,63 | 42,52 | 45,47 | 26,74 | 54,35 | 81,08 | 79,00 | 85,00 | 72,00 | 79,27 |
| 2020-12-28 | 30,50 | 38,11 | 41,83 | 46,20 | 39,16 | 30,48 | 40,09 | 70,57 | 63,00 | 64,00 | 74,00 | 67,89 |
| 2021-01-04 | 39,14 | 36,41 | 40,19 | 46,42 | 40,54 | 34,22 | 42,63 | 76,85 | 73,00 | 90,00 | 81,00 | 80,21 |
| 2021-01-10 | 23,44 | 28,72 | 33,82 | 42,52 | 32,12 | 39,16 | 16,37 | 55,53 | 56,00 | 73,00 | 69,00 | 63,38 |
| 2021-01-18 | 21,64 | 27,56 | 9,6 | 37,70 | 24,13 | 48,91 | 22,51 | 71,41 | 64,00 | 56,00 | 76,00 | 66,85 |
| 2021-01-25 | | 21,30 | 24,38 | 38,29 | 27,99 | | | | 61,00 | 71,00 | 71,00 | 67,67 |
| 2021-02-01 | | 32,51 | 26,58 | 28,60 | 29,23 | | | | 67,00 | 78,00 | 71,00 | 72,00 |
| 2021-02-08 | | 25,48 | 21,49 | 24,32 | 23,76 | | | | 73,00 | 64,00 | 69,00 | 68,67 |

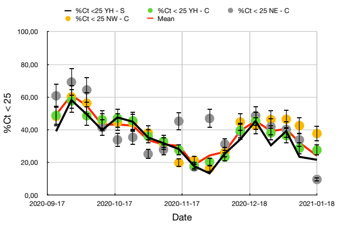

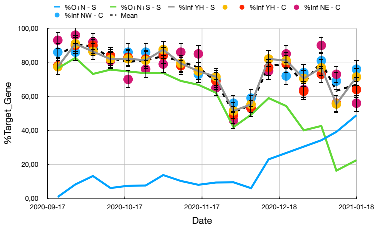

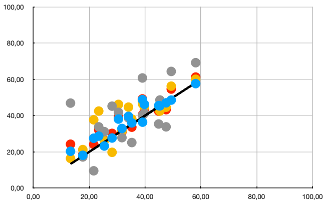

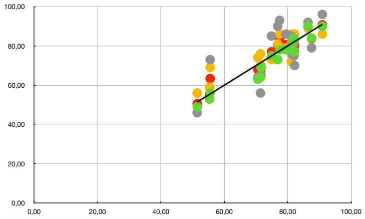

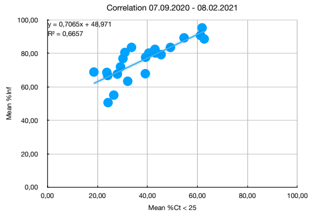

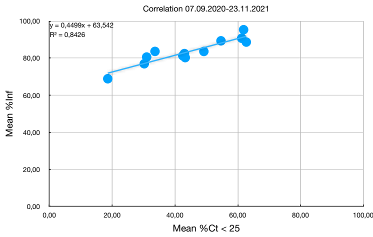

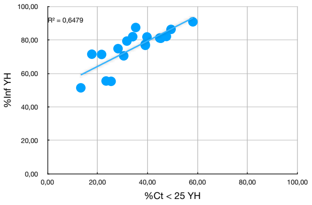

Steven

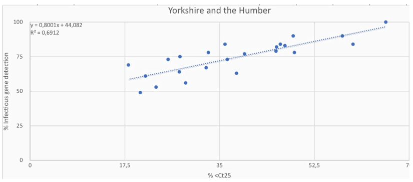

Charlotte

### Midlands

Table 1

| Date | %Ct < 25 WM - S | | %Ct < 25 WM - C | %Ct < 25 EM - C | %Ct < 25 SW - C | Mean | | %O+N - S | %O+N+S - S | %Inf WM - S | | %Inf WM - C | %Inf EM - C | %Inf SW - C | Mean |
|---|---|---|---|---|---|---|---|---|---|---|---|---|---|---|---|
| 2020-09-07 | | | 49,39 | 72,28 | 8,51 | 43,39 | | | | | | 50,00 | 71,00 | | 60,50 |
| 2020-09-14 | | | 51,25 | 24,04 | 29,81 | 35,03 | | | | | | 85,00 | 67,00 | 67,00 | 73,00 |
| 2020-09-21 | 50,43 | 50,43 | 48,01 | 25,79 | 27,84 | 38,02 | | 11,66 | 82,46 | 94,12 | 94,12 | 91,00 | 60,00 | 43,00 | 72,03 |
| 2020-09-28 | 45,22 | 45,22 | 44,48 | 34,13 | 31,62 | 38,86 | | 8,12 | 81,33 | 89,45 | 89,45 | 89,00 | 75,00 | 84,00 | 84,36 |
| 2020-10-05 | 51,30 | 51,30 | 47,32 | 62,28 | 31,36 | 48,07 | | 6,99 | 77,80 | 84,80 | 84,80 | 83,00 | 87,00 | 65,00 | 79,95 |
| 2020-10-12 | 46,96 | 46,96 | 48,14 | 45,24 | 27,91 | 42,06 | | 3,46 | 80,28 | 83,73 | 83,73 | 84,00 | 81,00 | 83,00 | 82,93 |
| 2020-10-19 | 40,87 | 40,87 | 41,29 | 41,80 | 33,27 | 39,31 | | 5,93 | 73,15 | 79,08 | 79,08 | 76,00 | 76,00 | 77,00 | 77,02 |
| 2020-10-26 | 48,70 | 48,70 | 47,00 | 40,67 | 41,46 | 44,46 | | 6,01 | 81,62 | 87,62 | 87,62 | 83,00 | 82,00 | 86,00 | 84,66 |
| 2020-11-02 | 34,78 | 34,78 | 38,07 | 39,30 | 39,02 | 37,79 | | 7,27 | 81,69 | 88,96 | 88,96 | 84,00 | 80,00 | 87,00 | 84,99 |
| 2020-11-09 | 36,52 | 36,52 | 35,69 | 36,68 | 39,05 | 36,99 | | 12,15 | 66,16 | 78,30 | 78,30 | 74,00 | 74,00 | 74,00 | 75,08 |
| 2020-11-16 | 29,57 | 29,57 | 28,01 | 32,57 | 32,38 | 30,63 | | 8,62 | 74,63 | 83,25 | 83,25 | 78,00 | 77,00 | 82,00 | 80,06 |
| 2020-11-23 | 34,78 | 34,78 | 33,14 | 22,79 | 26,63 | 29,34 | | 8,69 | 74,70 | 83,39 | 83,39 | 78,00 | 61,00 | 78,00 | 75,10 |
| 2020-11-30 | 11,30 | 11,30 | 11,07 | 22,37 | 34,13 | 19,72 | | 14,76 | 60,37 | 75,12 | 75,12 | 71,00 | 67,00 | 69,00 | 70,53 |
| 2020-12-07 | 26,96 | 26,96 | 24,39 | 26,29 | 11,62 | 22,31 | | 13,63 | 48,43 | 62,06 | 62,06 | 62,00 | 73,00 | 48,00 | 61,26 |
| 2020-12-14 | 38,26 | 38,26 | 39,56 | 38,64 | 47,64 | 41,03 | | 23,30 | 54,51 | 77,81 | 77,81 | 77,00 | 78,00 | 70,00 | 75,70 |
| 2020-12-21 | 46,96 | 46,96 | 40,41 | 51,74 | 30,57 | 42,42 | | 47,38 | 34,17 | 81,55 | 81,55 | 80,00 | 84,00 | 56,00 | 75,39 |
| 2020-12-28 | 48,70 | 48,70 | 45,72 | 35,20 | 29,32 | 39,73 | | 52,25 | 35,45 | 87,69 | 87,69 | 87,00 | 78,00 | 64,00 | 79,17 |
| 2021-01-04 | 61,74 | 61,74 | 59,79 | 30,73 | 47,18 | 49,86 | | 79,92 | 16,31 | 96,23 | 96,23 | 92,00 | 79,00 | 78,00 | 86,31 |
| 2021-01-10 | 39,13 | 39,13 | 36,14 | 24,57 | 30,45 | 32,57 | | 66,79 | 15,18 | 81,97 | 81,97 | 80,00 | 65,00 | 79,00 | 76,49 |
| 2021-01-18 | 20,87 | 20,87 | 30,60 | 25,56 | 18,38 | 23,85 | | 62,06 | 15,25 | 77,31 | 77,31 | 70,00 | 69,00 | 56,00 | 68,08 |
| 2021-01-25 | | | 28,82 | 33,53 | 25,91 | 29,42 | | | | | | 71,00 | 62,00 | 61,00 | 64,67 |
| 2021-02-01 | | | 26,26 | 29,97 | 9,92 | 22,05 | | | | | | 68,00 | 70,00 | 59,00 | 65,67 |
| 2021-02-08 | | | 28,78 | 20,47 | 19,47 | 22,91 | | | | | | 75,00 | 66,00 | 63,00 | 68,00 |

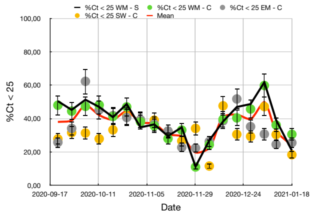

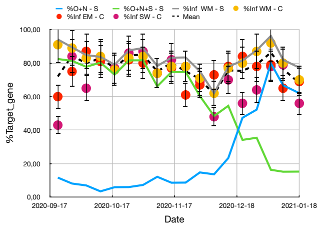

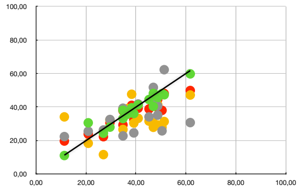

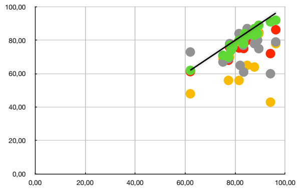

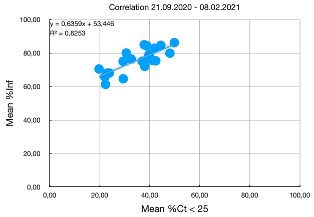

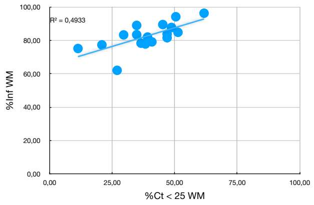

Steven

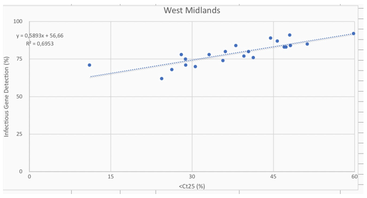

Charlotte

### LSE

Table 1

| Date | %Ct < 25 L - S | | %Ct < 25 L - C | %Ct < 25 SE - C | %Ct < 25 EE - C | Mean | | %O+N - S | %O+N+S - S | %Inf L - S | | %Inf L - C | %inf SE - C | %Inf EE - C | Mean |
|---|---|---|---|---|---|---|---|---|---|---|---|---|---|---|---|
| | 0 | 0 | | | | | | | | | | | | | |
| 2020-09-07 | | | 41,40 | 22,81 | 31,12 | 31,78 | | | | | | 81,00 | 100,00 | 85,00 | 88,67 |
| 2020-09-14 | | | 35,78 | 34,42 | 36,77 | 35,66 | | | | | | 78,00 | 60,00 | 84,00 | 74,00 |
| 2020-09-21 | 36,37 | 36,37 | 41,07 | 29,77 | 38,16 | 36,34 | | 13,28 | 79,95 | 93,23 | 93,23 | 91,00 | 60,00 | 73,00 | 79,31 |
| 2020-09-28 | 36,05 | 36,05 | 34,84 | 35,30 | 38,51 | 36,17 | | 17,66 | 68,77 | 86,43 | 86,43 | 85,00 | 76,00 | 75,00 | 80,61 |
| 2020-10-05 | 29,85 | 29,85 | 36,18 | 19,20 | 28,8 | 28,51 | | 14,26 | 52,04 | 66,30 | 66,30 | 67,00 | 43,00 | 35,00 | 66,65 |
| 2020-10-12 | 44,53 | 44,53 | 42,16 | 34,16 | 36,45 | 39,32 | | 11,98 | 67,52 | 79,50 | 79,50 | 80,00 | 81,00 | 78,00 | 79,63 |
| 2020-10-19 | 33,34 | 33,34 | 33,72 | 43,62 | 40,08 | 37,69 | | 14,13 | 54,13 | 68,25 | 68,25 | 72,00 | 82,00 | 73,00 | 73,81 |
| 2020-10-26 | 36,34 | 36,34 | 36,51 | 52,67 | 40,66 | 41,54 | | 17,40 | 66,28 | 83,68 | 83,68 | 82,00 | 79,00 | 78,00 | 80,67 |
| 2020-11-02 | 35,15 | 35,15 | 36,01 | 38,88 | 32,92 | 35,74 | | 11,78 | 60,66 | 72,44 | 72,44 | 75,00 | 81,00 | 77,00 | 76,36 |
| 2020-11-09 | 35,63 | 35,63 | 37,12 | 39,08 | 37,83 | 37,41 | | 17,27 | 61,71 | 78,98 | 78,98 | 82,00 | 79,00 | 75,00 | 78,74 |
| 2020-11-16 | 28,63 | 28,63 | 31,72 | 35,71 | 30,65 | 31,68 | | 21,64 | 57,20 | 78,84 | 78,84 | 84,00 | 85,00 | 85,00 | 83,21 |
| 2020-11-23 | 33,28 | 33,28 | 38,48 | 33,13 | 34,31 | 34,80 | | 33,80 | 52,70 | 86,49 | 86,49 | 89,00 | 82,00 | 75,00 | 83,12 |
| 2020-11-30 | 42,11 | 42,11 | 46,70 | 35,90 | 36,95 | 40,42 | | 37,07 | 38,18 | 75,25 | 75,25 | 83,00 | 73,00 | 83,00 | 78,56 |
| 2020-12-07 | 44,28 | 44,28 | 50,13 | 35,07 | 50,62 | 45,02 | | 53,67 | 25,89 | 79,57 | 79,57 | 87,00 | 67,00 | 92,00 | 81,39 |
| 2020-12-13 | 55,61 | 55,61 | 61,21 | 60,98 | 60,92 | 59,68 | | 68,05 | 16,95 | 85,00 | 85,00 | 90,00 | 88,00 | 90,00 | 88,25 |
| 2020-12-20 | 57,77 | 57,77 | 65,58 | 59,00 | 61,83 | 61,04 | | 75,76 | 9,10 | 84,86 | 84,86 | 93,00 | 92,00 | 90,00 | 89,97 |
| 2020-12-28 | 42,41 | 42,41 | 56,64 | 49,92 | 56,03 | 51,25 | | 73,46 | 5,70 | 79,16 | 79,16 | 90,00 | 83,00 | 94,00 | 86,54 |
| 2021-01-04 | 29,57 | 29,57 | 34,91 | 44,50 | 46,48 | 38,86 | | 76,74 | 4,53 | 81,26 | 81,26 | 83,00 | 83,00 | 89,00 | 84,07 |
| 2021-01-11 | 24,22 | 24,22 | 29,40 | 31,03 | 24,92 | 27,39 | | 66,67 | 3,35 | 70,02 | 70,02 | 78,00 | 74,00 | 67,00 | 72,25 |
| 2021-01-18 | 20,52 | 20,52 | 25,03 | 23,86 | 28,31 | 24,43 | | 64,39 | 2,17 | 66,56 | 66,56 | 69,00 | 67,00 | 64,00 | 66,64 |
| 2021-01-25 | | | 19,76 | 19,82 | 26,01 | 21,86 | | | | | | 64,00 | 61,00 | 64,00 | 63,00 |
| 2021-02-01 | | | 19,86 | 19,08 | 9,52 | 16,15 | | | | | | 57,00 | 53,00 | 51,00 | 53,67 |
| 2021-02-08 | | | 20,91 | 24,70 | 24,71 | 23,44 | | | | | | 58,00 | 63,00 | 59,00 | 60,00 |
| | 100 | 100 | | | | | | | | | | | | | |

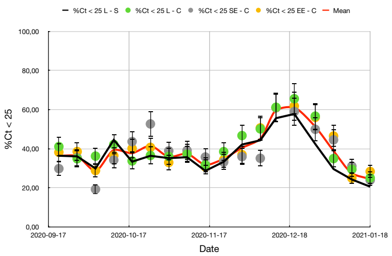

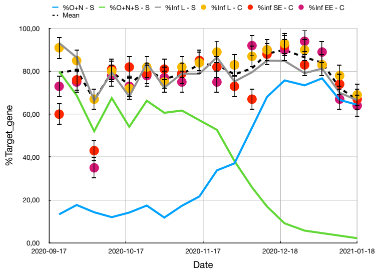

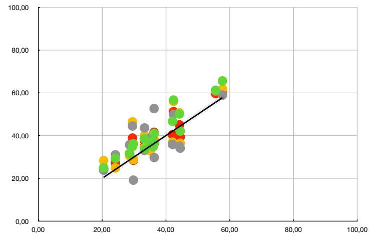

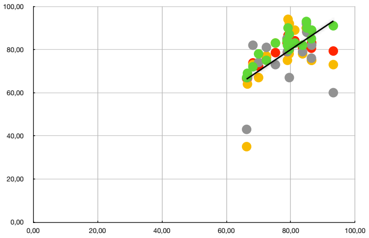

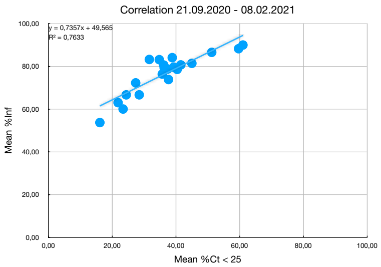

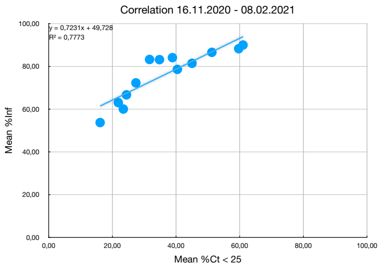

Charlotte

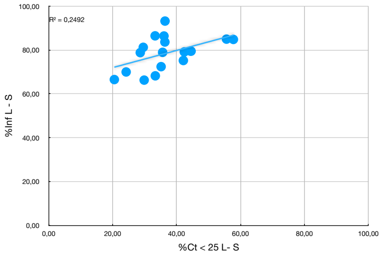

Steven

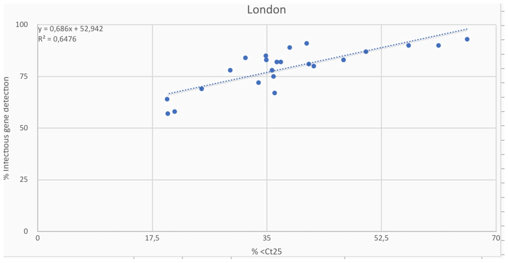
