## Supplementary material for "An Analysis of PCR Ct Scores and Distributions from the ONS Community Infection Survey during the COVID Second Wave in the UK": P(Ct) datasets and graphs

### North

Table 1

| | Date | YH - Ct Pk hi | | YH - Ct Pk lo | | NW - Ct Pk hi | NW - Ct Pk lo | NE - Ct Pk hi | NE - Ct Pk lo | Mean Ct Pk hi | Mean Ct Pk lo |
|---|---|---|---|---|---|---|---|---|---|---|---|
| | 2020-09-07 | | | | | | | | | | |
| | 2020-09-14 | | | | | | | | | | |
| | 2020-09-21 | 28,45 | 28,45 | 19,08 | | 29,71 | 17,77 | 24,20 | | 27,46 | 18,42 |
| | 2020-09-28 | 30,66 | 30,66 | 18,16 | | 30,03 | 19,93 | 27,34 | 19,26 | 29,34 | 19,12 |
| | 2020-10-05 | 31,30 | 31,30 | 18,18 | | 30,05 | 17,81 | 25,87 | 16,63 | 29,07 | 17,54 |
| | 2020-10-12 | 32,26 | 32,26 | | | 31,91 | | 31,60 | 22,66 | 31,92 | 22,66 |
| | 2020-10-19 | 32,90 | 32,90 | 17,90 | | 31,01 | 18,76 | 32,72 | 18,30 | 32,21 | 18,32 |
| | 2020-10-26 | 32,29 | 32,29 | 18,54 | | 30,72 | | 32,98 | 23,46 | 32,00 | 21,00 |
| | 2020-11-02 | 32,62 | 32,62 | 20,44 | | 31,96 | 18,79 | 32,08 | 19,39 | 32,22 | 19,54 |
| | 2020-11-09 | 32,96 | 32,96 | 19,52 | | 32,28 | | 31,76 | 21,08 | 32,33 | 20,30 |
| | 2020-11-16 | 34,22 | 34,22 | 20,79 | | 33,22 | | 32,30 | 18,45 | 33,25 | 19,62 |
| | 2020-11-23 | 34,87 | 34,87 | | | 32,93 | | 33,13 | 25,00 | 33,64 | 25,00 |
| | 2020-11-30 | 35,51 | 35,51 | | | 34,48 | | 33,67 | 30,00 | 34,55 | 30,00 |
| | 2020-12-07 | 35,22 | 35,22 | 19,59 | | 33,58 | 19,80 | 32,77 | | 33,86 | 19,70 |
| | 2020-12-13 | 33,05 | 33,05 | 19,61 | | 32,07 | 17,07 | 30,72 | 16,87 | 31,95 | 17,85 |
| | 2020-12-20 | 34,32 | 34,32 | 18,38 | | 33,00 | 18,31 | 32,70 | 14,82 | 33,34 | 17,17 |
| | 2020-12-28 | 35,27 | 35,27 | | | 33,94 | 17,72 | 33,82 | 17,96 | 34,35 | 17,84 |
| | 2021-01-04 | 33,73 | 33,73 | 21,85 | | 33,35 | | 27,73 | | 31,60 | 21,85 |
| | 2021-01-11 | 33,74 | 33,74 | 21,87 | | 33,98 | | 32,89 | 19,05 | 33,54 | 20,46 |
| | 2021-01-18 | 34,08 | 34,08 | 23,45 | | 33,38 | 19,61 | 33,72 | | 33,73 | 21,53 |
| | 2021-01-25 | | | | | | | | | 32,24 | 20,44 |
| | 2021-02-01 | | | | | | | | | | |
| | 2021-02-08 | | | | | | | | | | |

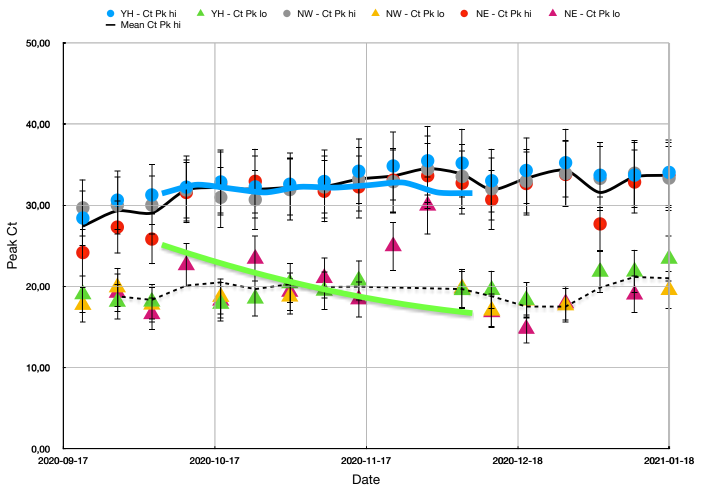

### Midlands

Table 1

| Date | WM - Ct Pk hi | | WM - Ct Pk lo | | EM - Ct Pk hi | EM - Ct Pk lo | SW - Ct Pk hi | SW -Ct Pk lo | Mean Ct Pk hi | Mean Ct Pk lo | |
|---|---|---|---|---|---|---|---|---|---|---|---|
| 2020-09-07 | | | | | | | | | | | |
| 2020-09-14 | | | | | | | | | | | |
| 2020-09-21 | 28,45 | | 14,68 | | 30,59 | | 34,98 | 28,42 | 31,34 | 21,55 | 21,55 |
| 2020-09-28 | 30,88 | | 18,03 | | 30,87 | 20,67 | 31,22 | 20,59 | 30,99 | 19,76 | 19,76 |
| 2020-10-05 | 33,01 | | 17,40 | | 31,73 | 17,95 | 34,32 | 24,95 | 33,02 | 20,10 | 20,10 |
| 2020-10-12 | 27,48 | | | | 32,61 | 23,63 | 30,24 | | 30,11 | 23,63 | 23,63 |
| 2020-10-19 | 30,22 | | 18,89 | | 32,30 | 18,21 | 30,85 | | 31,12 | 18,55 | 18,55 |
| 2020-10-26 | 31,73 | | 18,57 | | 32,28 | 23,60 | 31,77 | 23,33 | 31,93 | 21,83 | 21,83 |
| 2020-11-02 | 31,71 | | 21,31 | | 32,56 | | 32,69 | | 32,32 | 21,31 | 21,31 |
| 2020-11-09 | 32,31 | | | | 32,85 | 18,46 | 33,61 | 19,54 | 32,92 | 19,00 | 19,00 |
| 2020-11-16 | 33,21 | | | | 34,03 | | 31,09 | 19,53 | 32,77 | 19,53 | 19,53 |
| 2020-11-23 | 30,43 | | | | 34,01 | | 33,57 | 27,94 | 32,67 | 27,94 | |
| 2020-11-30 | 32,56 | | | | 33,99 | 23,81 | 33,24 | 21,99 | 33,26 | 22,90 | 22,90 |
| 2020-12-07 | 33,46 | | 21,83 | | 33,07 | 27,39 | 34,16 | 31,34 | 33,56 | 26,85 | |
| 2020-12-13 | 31,91 | | | | 32,46 | 17,47 | 33,83 | 17,26 | 32,73 | 17,37 | 17,37 |
| 2020-12-20 | 31,89 | | 18,43 | | 33,04 | 15,36 | 33,81 | 27,24 | 32,91 | 20,34 | 20,34 |
| 2020-12-28 | 32,49 | | 17,79 | | 33,02 | 18,94 | 34,73 | | 33,41 | 18,37 | 18,37 |
| 2021-01-04 | | | 22,67 | | 33,30 | 17,42 | 32,52 | 16,58 | 32,91 | 18,89 | 18,89 |
| 2021-01-11 | 32,45 | | 18,68 | | 33,29 | | 33,44 | | 33,06 | 18,68 | 18,68 |
| 2021-01-18 | 33,66 | | | | 30,59 | | 34,36 | | 32,87 | | |
| 2021-01-25 | | | | | | | | | 32,44 | 20,98 | 20,12 |
| 2021-02-01 | | | | | | | | | | | |
| 2021-02-08 | | | | | | | | | | | |

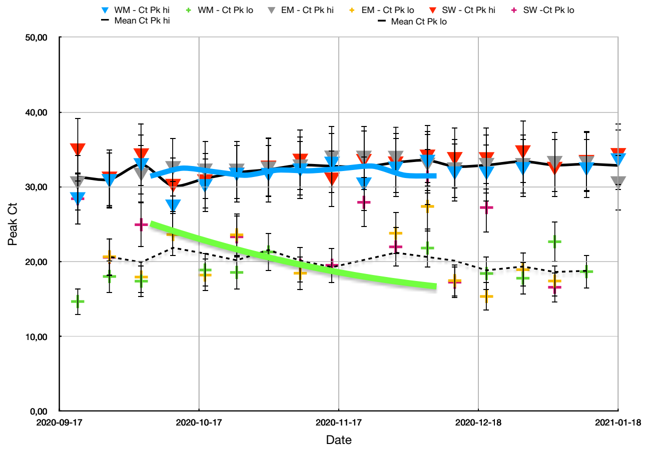

### LSE

Table 1

| | Date | L - Ct Pk hi | | L - Ct Pk lo | | SE - Ct Pk hi | Se - Ct Pk lo | EE - Ct Pk hi | EE - Ct Pk lo | Mean Ct Pk hi | Mean Ct Pk lo |
|---|---|---|---|---|---|---|---|---|---|---|---|
| | 2020-09-07 | | | | | | | | | | |
| | 2020-09-14 | | | | | | | | | | |
| | 2020-09-21 | 29,42 | | 21,46 | | 35,95 | 25,64 | 33,13 | 24,06 | 32,83 | 23,72 |
| | 2020-09-28 | 32,82 | | 25,78 | | 33,47 | | 34,38 | 22,50 | 33,56 | 24,14 |
| | 2020-10-05 | 34,70 | | 20,61 | | 34,43 | 25,68 | 34,38 | 22,19 | 34,50 | 22,82 |
| | 2020-10-12 | 34,12 | | 19,42 | | 33,20 | 25,70 | 33,13 | 20,31 | 33,48 | 21,81 |
| | 2020-10-19 | 35,07 | | 21,60 | | 32,59 | 19,77 | 33,44 | 20,31 | 33,70 | 20,56 |
| | 2020-10-26 | 33,27 | | | | 28,23 | 17,30 | 32,81 | 19,38 | 31,44 | 18,34 |
| | 2020-11-02 | 33,31 | | | | 30,13 | | 33,44 | | 32,29 | |
| | 2020-11-09 | 33,65 | | 20,18 | | 33,58 | 18,27 | 34,06 | 19,06 | 33,76 | 19,17 |
| | 2020-11-16 | 33,69 | | | | 32,66 | 20,47 | 32,50 | 22,19 | 32,95 | 21,33 |
| | 2020-11-23 | 30,66 | | 19,94 | | 33,31 | 20,18 | 33,13 | 20,63 | 32,36 | 20,25 |
| | 2020-11-30 | 33,76 | | 19,67 | | 34,26 | 23,95 | 31,56 | 20,94 | 33,19 | 21,52 |
| | 2020-12-07 | 32,57 | | 17,88 | | 34,28 | 20,22 | 30,63 | | 32,49 | 19,05 |
| | 2020-12-13 | | | 16,07 | | 32,11 | 16,49 | 31,88 | 15,63 | 31,99 | 16,06 |
| | 2020-12-20 | | | 18,25 | | 31,82 | 17,13 | 31,88 | 16,25 | 31,85 | 17,21 |
| | 2020-12-28 | 32,98 | | | | 33,71 | 18,40 | 28,13 | 18,13 | 31,61 | 18,26 |
| | 2021-01-04 | 33,02 | | | | 30,60 | | 30,63 | 17,81 | 31,42 | 17,81 |
| | 2021-01-11 | 35,20 | | 21,42 | | 34,69 | | 34,69 | | 34,86 | 21,42 |
| | 2021-01-18 | 34,93 | | 21,76 | | 35,02 | | 34,38 | | 34,77 | 21,76 |
| | 2021-01-25 | | | | | | | | | 32,95 | 20,31 |
| | 2021-02-01 | | | | | | | | | | |
| | 2021-02-08 | | | | | | | | | | |

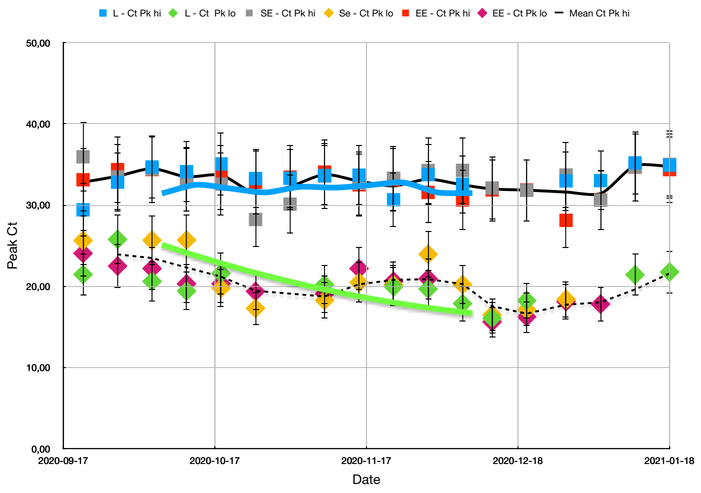

### Gene Targets

Table 1

| | Date | O+N Ct Pk hi | O+N Ct Pk lo | O+N+S Ct Pk hi | O+N+S Ct lo |
|---|---|---|---|---|---|
| | 2020-10-07 | 31,44 | | 29,08 | 16,67 |
| | 2020-10-14 | 32,51 | | 28,09 | 16,90 |
| | 2020-10-21 | 32,04 | 22,46 | 27,92 | 16,73 |
| | 2020-10-28 | 31,56 | | 28,54 | 16,55 |
| | 2020-11-04 | 32,25 | | 28,35 | 17,18 |
| | 2020-11-11 | 32,15 | | 28,59 | 15,80 |
| | 2020-11-18 | 32,46 | 18,66 | 29,60 | 18,42 |
| | 2020-11-25 | 32,78 | 17,80 | 29,02 | 17,84 |
| | 2020-12-02 | 31,56 | 16,94 | 29,64 | 16,47 |
| | 2020-12-09 | 31,48 | 16,85 | 28,26 | 15,88 |
| | | 32,02 | 18,54 | | |

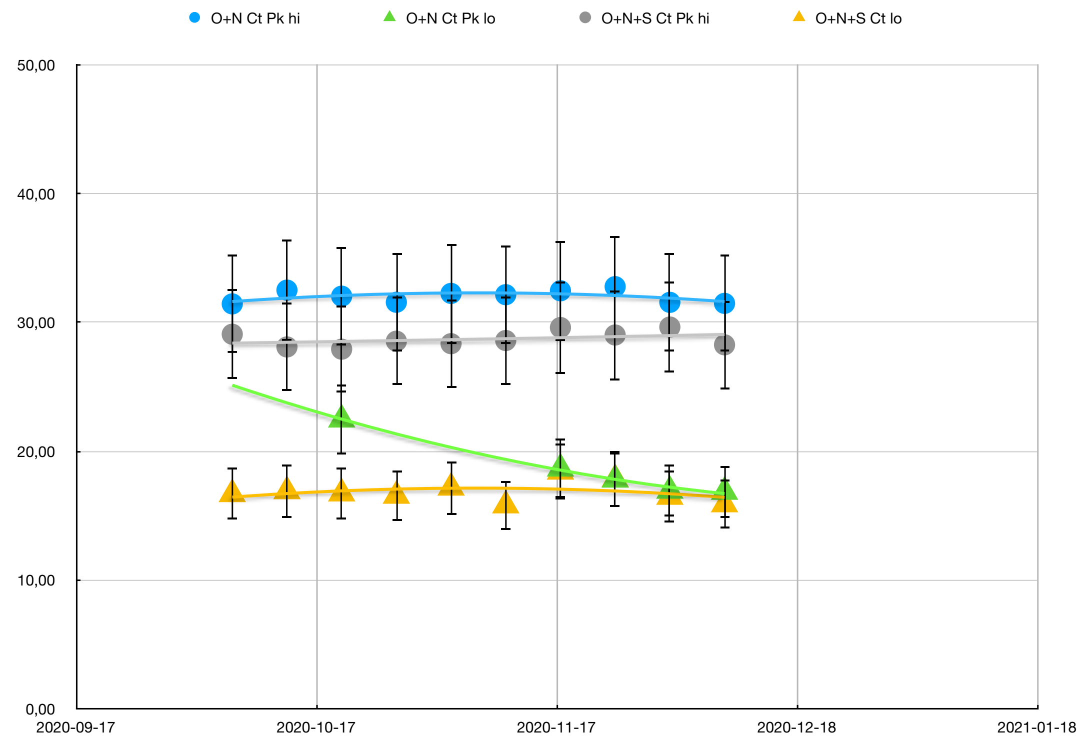
